# Microhaplotype deep sequencing assays to capture *Plasmodium vivax* infection lineages

**DOI:** 10.1101/2024.10.14.24315131

**Authors:** Mariana Kleinecke, Angela Rumaseb, Edwin Sutanto, Hidayat Trimarsanto, Kian Soon Hoon, Ashley Osborne, Paulo Manrique, Trent Peters, David Hawkes, Ernest Diez Benavente, Georgia Whitton, Sasha V Siegel, Richard D Pearson, Roberto Amato, Anjana Rai, Nguyen Thanh Thuy Nhien, Nguyen Hoang Chau, Ashenafi Assefa, Tamiru S Degaga, Dagimawie Tadesse Abate, Awab Ghulam Rahim, Ayodhia Pitaloka Pasaribu, Inge Sutanto, Mohammad Shafiul Alam, Zuleima Pava, Tatiana Lopera- Mesa, Diego Echeverry, Tim William, Nicholas M Anstey, Matthew J Grigg, Nicholas Day, Nicholas J. White, Dominic P Kwiatkowski, Rintis Noviyanti, Daniel Neafsey, Ric N Price, Sarah Auburn

**Affiliations:** Menzies School of Health Research and Charles Darwin University, Darwin, Australia; Exeins Health Initiative, South Jakarta, Indonesia; Harvard T.H. Chan School of Public Health, USA; Broad Institute, USA; Australian Genome Research Facility, Australia; Laboratory of Experimental Cardiology, Department of Cardiology, University Medical Center Utrecht, Utrecht, the Netherlands; Wellcome Sanger Institute, Hinxton, UK; Oxford University Clinical Research Unit, Hospital for Tropical Diseases, Ho Chi Minh City, Vietnam; Ethiopian Public Health Institute, Addis Ababa, Ethiopia; College of Medicine & Health Sciences, Arba Minch University, Arba Minch, Ethiopia; Nangarhar Medical Faculty, Nangarhar University, Ministry of Higher Education, Jalalabad, Afghanistan; Universitas Sumatera Utara, Medan, Indonesia; Faculty of Medicine, University of Indonesia, Jakarta, Indonesia; Infectious Diseases Division, International Centre for Diarrhoeal Disease Research, Bangladesh (icddr,b), Dhaka, Bangladesh; Universidad de Antioquia, Colombia; Departamento de Microbiología, Facultad de Salud, Universidad del Valle, Cali, Colombia; Queen Elizabeth Hospital, Malaysia; Centre for Tropical Medicine and Global Health, Nuffield Department of Medicine, University of Oxford, Oxford, United Kingdom; Mahidol-Oxford Tropical Medicine Research Unit, Mahidol University, Bangkok, Thailand; Eijkman Molecular Biology Research Center, National Research and Innovation Agency, Cibinong, Indonesia

**Author notes:** Deceased (Dominic P Kwiatkowski). Corresponding author: A/Prof Sarah Auburn, Menzies School of Health Research, PO Box 41096, Casuarina, Darwin, NT 0811, Australia.

**Keywords:** Plasmodium vivax, malaria, microhaplotype, relatedness, identity-by-descent, recurrence, relapse, amplicon sequencing

## Abstract

The elimination of *Plasmodium vivax* is challenged by dormant liver stages (hypnozoites) that can reactivate months after initial infection resulting in relapses that enhance transmission. Relapsing infections confound antimalarial clinical efficacy trials due to the inability to distinguish between recurrences arising from blood-stage treatment failure (recrudescence), reinfection or relapse. Genetic relatedness of paired parasite isolates, measured by identity-by-descent (IBD), can provide important information on whether individuals have had single or multiple mosquito inoculations, thus informing on recurrence origin. We developed a high-throughput amplicon sequencing assay comprising 93 multi-SNP (microhaplotype) *P. vivax* markers to determine IBD between *P. vivax* clinical isolates. The assay was evaluated in 659 independent infections from the Asia-Pacific and Horn of Africa, including 108 pairs of infections from a randomized controlled trial (RCT). A bioinformatics pipeline (vivaxGEN-MicroHaps) was established to support data processing. Simulations using *paneljudge* demonstrated low error in pairwise IBD estimation in all countries assessed (all RMSE <0.12) and IBD-based networks illustrated strong clustering by geography. IBD analysis in the RCT demonstrated a higher frequency of suspected relapses or recrudescence in patients treated with primaquine regimens (0.84) compared to those without primaquine (0.60). Our results demonstrate the potential to derive information on *P. vivax* IBD using amplicon sequencing, that informs policy-relevant treatment and transmission dynamics.

## Introduction

Outside of sub-Saharan Africa, *Plasmodium vivax* is becoming the predominant cause of malaria ^1^. The control and elimination of *P. vivax* is confounded by the parasite’s ability to form dormant liver stages (hypnozoites) that can reactivate weeks to months after initial infection, causing recurrent episodes of malaria known as relapses ^2^. The radical cure of vivax malaria requires treatment with both a schizontocidal antimalarial (chloroquine (CQ) or artemisinin combination therapy) combined with a hypnozoiticidal agent (primaquine or tafenoquine)^1^. *P. vivax* resistant to chloroquine (CQR) has been documented in several endemic countries^3,4^, but surveillance of the emergence and spread of CQR is undermined by a lack of reliable molecular markers ^2,5^. Although chloroquine efficacy can be defined using clinical efficacy trials to determine the risk of recurrent parasitaemia within 28 to 42 days of treatment, estimates are confounded by an inability to distinguish whether recurrent infections are due to schizontocidal treatment failure (recrudescence), reactivation of hypnozoites (relapses), or a new mosquito inoculation (reinfection) ^6,7^. In the case a resistant infection is detected, traditional surveillance tools, such as patient travel history, may also be limited in capacity to assess the origin or spread of the given strains within a community ^2^. Molecular approaches provide great potential to address these knowledge gaps ^6,8^.

Parasite genotyping of infection pairs (pre- and post-treatment), is well established for interpreting antimalarial clinical efficacy for *P. falciparum*, to distinguish between recrudescence (same/homologous pairs) and reinfection (different/heterologous pairs) ^9^. However, a similar approach to defining clinical efficacy for *P. vivax* infection is undermined by relapses that can be homologous or heterologous to the pre-treatment isolate ^10–12^. Genomic studies of *P. vivax* have revealed that a proportion of paired clinical isolates that are classified as heterologous using traditional genotyping methods, can share homology in large segments of the genome, inferring recent common ancestry^13,14^. These familial relationships can be defined using genetic information on identity-by-descent (IBD). Closely related paired infections (e.g., siblings, with ≥50% IBD) are more likely to come from a single mosquito inoculation and thus to be relapses rather than reinfections. Genome-wide data are ideal for quantifying IBD. However, this is not feasible in most malaria-endemic settings. Targeted next generation sequencing (NGS)-based methods, such as amplicon sequencing, offer an affordable and versatile alternative ^15^. Previously we have established a bioinformatic framework for retrieving genome-wide panels of *P. vivax* microhaplotype markers; short (200 bp) genomic segments comprising multiple high diversity single nucleotide polymorphisms (SNPs) ^8^. Using *in-silico* analyses, we demonstrated that ∼100 *P. vivax* microhaplotypes can reliably captured IBD between paired isolates and could be used to define spatio-temporal patterns of *P. vivax* infection^8^.

In this study, we advanced our previous *in-silico* work by designing an NGS-based amplicon sequencing assay for 98 *P. vivax* amplicons, including 93 microhaplotypes that capture IBD accurately in diverse endemic areas. Our rhAmpSeq library preparation protocol incorporates all markers in a single plex reaction and was able to generate high throughput, cost-effective, sensitive and specific data from global isolates. Standard population-level genetic metrics could be applied to the data to highlight potential use cases for National Malaria Control Programs (NMCPs) that will inform *P. vivax* treatment options and transmission dynamics.

## Results

### A rhAmpSeq multiplex of 93 microhaplotypes with high sensitivity and specificity

A two-step multiplex PCR-based library preparation assay was established using rhAmpSeq chemistry (Integrated DNA Technologies, IDT). The multiplex amplifies one species confirmation marker, four putative markers of drug resistance, and 93 microhaplotypes distributed relatively uniformly across the *P. vivax* genome (Supplementary Data 1; Supplementary Figure 1). To evaluate the assays technical and analytical performance, a total of 750 *P. vivax* infections were assessed, including 108 paired *P. vivax* infections from a randomized controlled trial, two *P. vivax* serial dilution panels, and 10 non-*P. vivax* control samples (Supplementary Data 2, Supplementary Figure 2).

The specificity of the assay was evaluated using a selection of 7 non-vivax *Plasmodium* spp. and 3 uninfected human DNA controls. None (0/10) of the control samples were permissible to successful read pair calling using *DADA2*. Examination of amplicon coverage revealed that 58 markers (60% [58/97] after excluding the mitochondrial species marker) exhibited amplicon coverage <0.9 in all negative controls which would not yield successful read-pairs (Supplementary Figure 3a). Amongst the 39 markers with coverage >0.9 in one or more negative controls, 37 (95%) had read counts <25 in all controls tested, with only 2 markers (markers 307891 and 313080) exceeding 25 reads and only in the *P. knowlesi* controls (Supplementary Figure 3b).

The assay sensitivity was assessed using serial dilutions of two *P. vivax*-infected patient blood samples (KV3 and KV5) under high sample multiplexing (library pooled across 384 samples), using both the standard PCR step 1 DNA input and reaction volumes (referred to as full chemistry with a 20 ul reaction volume) and halving the PCR step 1 DNA input and reaction volumes (referred to as half chemistry with a 10 ul reaction volume). Under full chemistry conditions, depths exceeding 100 reads per marker were observed in the 70 parasite/ul (mean 95-160 across triplicates) and 96 parasite/ul (mean 74-138 across triplicates) sample preparations, as well as in higher densities (Supplementary Figures 4a and c). Under half chemistry conditions, similar depths were observed in the 70 parasite/ul (mean 235-280 across triplicates) and 96 parasite/ul (mean 167-194 across triplicates) sample preparations and higher parasite densities (Supplementary Figures 4b and d).

Parasite density defined by microscopic blood film examination may not correlate directly with parasite DNA yield owing to the presence of multiple life cycle stages in *P. vivax* infections (schizonts, for example, having greater DNA quantity than ring stages). To address this, we evaluated the threshold cycle (Ct) values of the KV5 serial dilution using real-time PCR, targeting the *pvmtcox1* gene. Samples with a Ct <30 (KV5 96 parasite/ul) yielded a mean of ≥100 reads or more, while samples with a Ct <33 (KV5 9.6 parasite/ul) resulted in a mean yield of ≥9 reads (Supplementary Figure 5, Supplementary Table 1).

### Geographically diverse application potential

A total of 659 *P. vivax* isolates were used to evaluate amplification efficacy from 8 countries across the globe. Successful genotyping was defined as ≥25 reads for >80% of the 97 nuclear genome amplicons and could be achieved in 627 (95.1%) isolates. 500 (79.7%) independent (non-recurrent) samples were taken forward for evaluation of individual marker efficacy and country-specific amplification patterns. A minimum of 5 isolates were available from 6 countries: Afghanistan (n=156), Bangladesh (n=5), Colombia (n=5), Ethiopia (n=213), Indonesia (n=38) and Vietnam (n=83). Aside from 146 samples from East Shewa, Ethiopia, and 38 samples from Sumatra, Indonesia, data from all isolates was generated from a single 384-well run (run 3). One marker (419038) demonstrated poor amplification in all run 3 populations with both read (SNP-based) and read-pair (microhaplotype-based) analyses (Supplementary Data 3, Figure 1). Marker 419038 was therefore excluded from several downstream analyses (see Supplementary Figure 1 for usage). However, increasing the primer concentration for marker 419038 enhanced amplification in later runs, as demonstrated in the East Shewa and Sumatran samples from runs 5 and 6. Three markers (64721, 354590 and 466426) exhibited a large drop in read-pair counts in a moderate proportion of samples after denoising with *DADA2* (Figure 1b). Marker 354590 was most impacted by the *DADA2* denoising and was excluded from all downstream read-pair analyses. The remaining 93 markers exhibited >50% successful genotype calls in majority of populations assessed.

**Figure 1.**
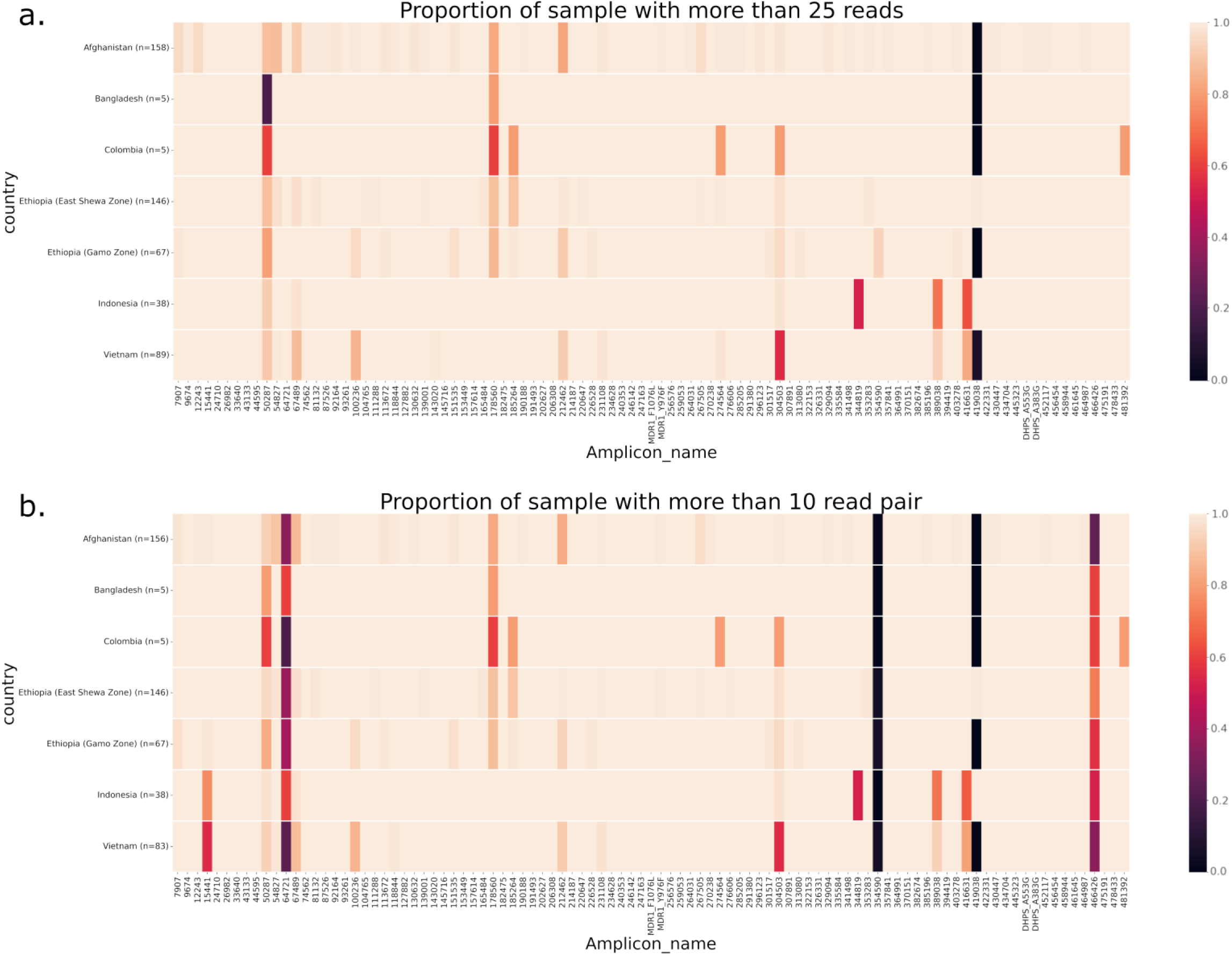
*P. vivax* proportion of samples with more than 25 reads or more than 10 read pairs by country. Heat maps illustrate the proportion of samples with more than 25 reads (panel a) and proportion of samples with more than 10 read pairs derived from *DADA2* (panel b) for each marker by country. Markers (x-axis) were ordered by chromosome and coordinate. All samples with genotype failures (<25 reads or <10 read pairs) are presented in black. Microhaplotype markers 64721, 354590, 419038 and 466426 displayed consistently low read-pair counts. Microhaplotype marker 354590 exhibits high individual read counts (panel a) but low read-pair counts (panel b) in all countries and was excluded from all downstream read pair analyses. Data are presented on a total of 500 independent (not including replicates) infections that passed genotyping. The data from Ethiopia are split into dried blood spot (DBS) from a clinical trial conducted in East Shewa Zone and whole blood (WB) extracts from a therapeutic efficacy survey conducted in Gamo Zone for comparative assessment. The DBS versus WB comparisons reveal similar read and read pair counts at all loci.

### Accuracy in P. vivax variant calling

Using a set of 83 isolates with both whole genome sequencing (WGS) and amplicon sequencing data, the accuracy of variant calling (applying the default 10% minor allele threshold) was confirmed at the 425 SNPs within the 93 microhaplotypes. Concordance was observed at 98.3% (32,352/32,923) of the genotype calls (Supplementary Table 2, Supplementary Data 4). Homozygous reference versus homozygous alternate allele discordances contributed 0.06% (19/32,923) of all calls. Most discordant calls reflected heterozygous versus homozygous calls (96.7% (552/571)), likely reflecting differences in the limit of detection of minor clones between the datasets. On further exploring the concordance after applying a 1% minor allele threshold to both datasets, a notable difference was more than doubling of heterozygous amplicon sequencing calls that were homozygous in the WGS dataset (from 225 calls with 10% threshold to 550 with 1% threshold; Supplementary Table 3); this trend likely reflects a greater limit of detection in the deep sequencing (amplicon) data.

### High potential to capture within-host infection complexity

Using a set of 83 isolates with both WGS and microhaplotype data, within-host infection complexity was explored using the *F*ws and effective MOI (eMOI). As illustrated in Figures 2a and b, there was a strong correlation between the genome-wide *F*ws and the microhaplotype-based eMOI in each geographic region assessed (Spearman’s rank correlation, rho=-0.700 *p*=2.389e-13). However, amongst 59 infections with *F*ws≥0.95 (traditionally used as a cut-off to define monoclonal infections), 5 isolates had >95% probability of being polyclonal according to MOIRE analysis of the microhaplotypes (eMOI range 1.08-1.93) (Supplementary Data 5). In contrast, none of the infections defined as polyclonal with genomic data (*F*ws<0.95) were monoclonal with the microhaplotypes (all 100% probability of polyclonality).

**Figure 2.**
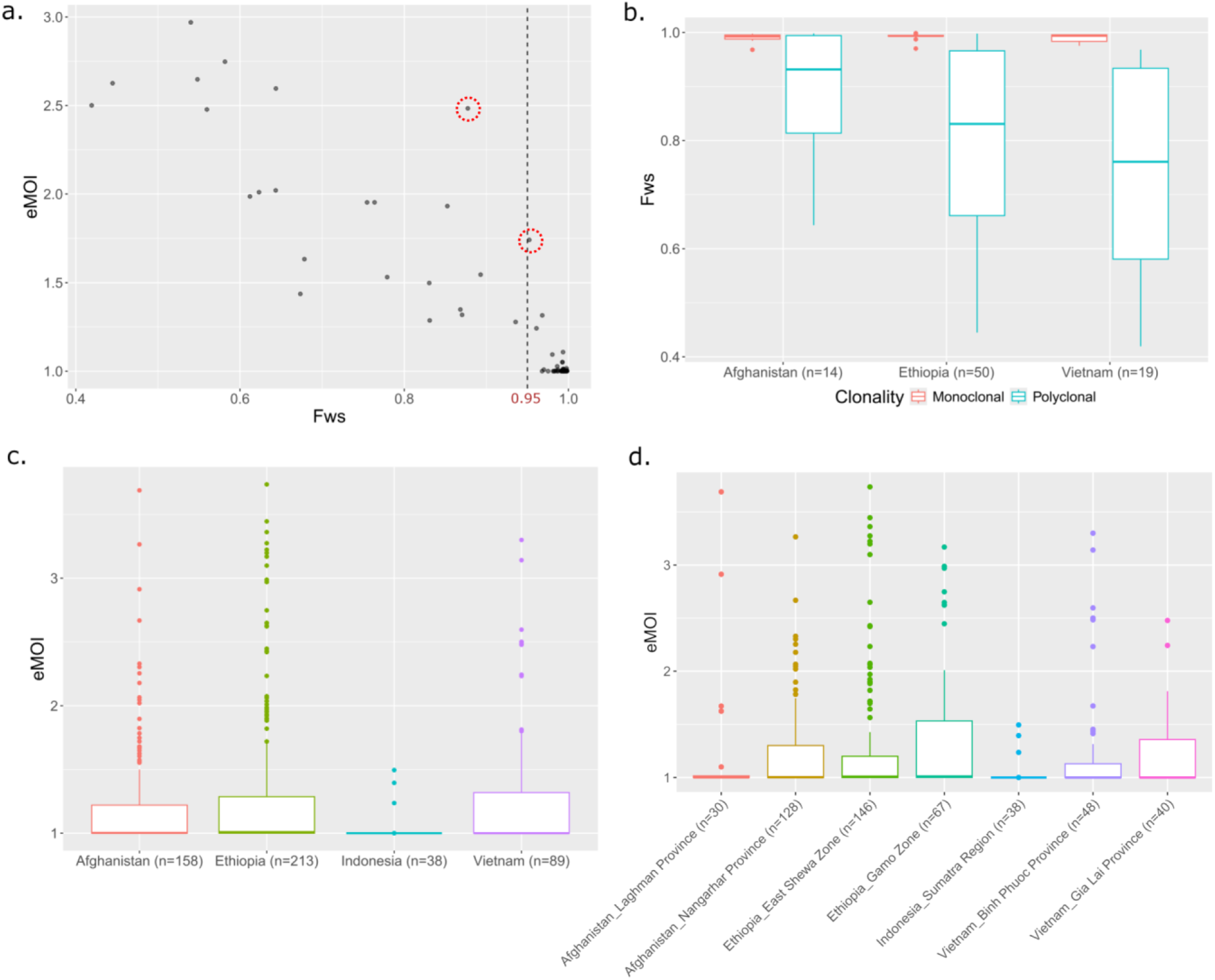
Microhaplotype-based within-host diversity trends. Panels a) and b) illustrate the level of concordance between genomic (as measured by the Fws) and microhaplotype (as measured by eMOI) data in estimation of within-host *P. vivax* diversity. The boxplots present the median, interquartile range and min and max value. Data are derived from 83 independent *P. vivax* cases. Overall, high concordance is observed between the two datasets. However, a few infections, including VN0001-321 and VN001_030 circled in panel 4a) display notably higher within-host diversity with microhaplotype versus whole genome sequence data. Panels c) and d) present the eMOI distributions at the country level (c) and provincial level (d), illustrating variation between sites, likely reflecting local endemicity levels. The data in panels c) and d) are based on 498 independent cases.

The eMOI distributions in Figures 2c and d illustrate trends in within-host diversity at the country and provincial level in a larger panel of 498 isolates from 4 populations which had over 30 isolates available: Afghanistan (n=158), Indonesia (n=38), Ethiopia (n=213) and Vietnam (n=89). The data reveals differences within-country in Afghanistan, where Laghman exhibited a trend (not significant) of lower eMOI than Nangahar (Wilcoxon rank sum test, *p*=0.324). The lowest eMOI distribution was observed in Sumatra, Indonesia, suggestive of low endemicity relative to the other sites.

### Effective IBD capture

To assess the ability of the microhaplotype panel to capture IBD accurately, we used *paneljudge* (an R package to judge the performance of a panel of genetic markers using simulated data) to simulate the relative mean square error (RMSE) in estimation of five pairwise IBD states: IBD=0 (unrelated infections), IBD=0.25 (half-siblings), IBD=0.5 (siblings), IBD=0.75 (highly related), and IBD=1 (identical clones). The simulations were run in the 4 major populations (Afghanistan, Ethiopia, Indonesia and Vietnam) using data from the 92 assayable microhaplotypes, revealing moderately high diversity (mean diversity range 0.44-0.65) and low RMSE (<0.12 for all pairwise IBD states) in each population (Supplementary Figure 6, Figure 3).

**Figure 3.**
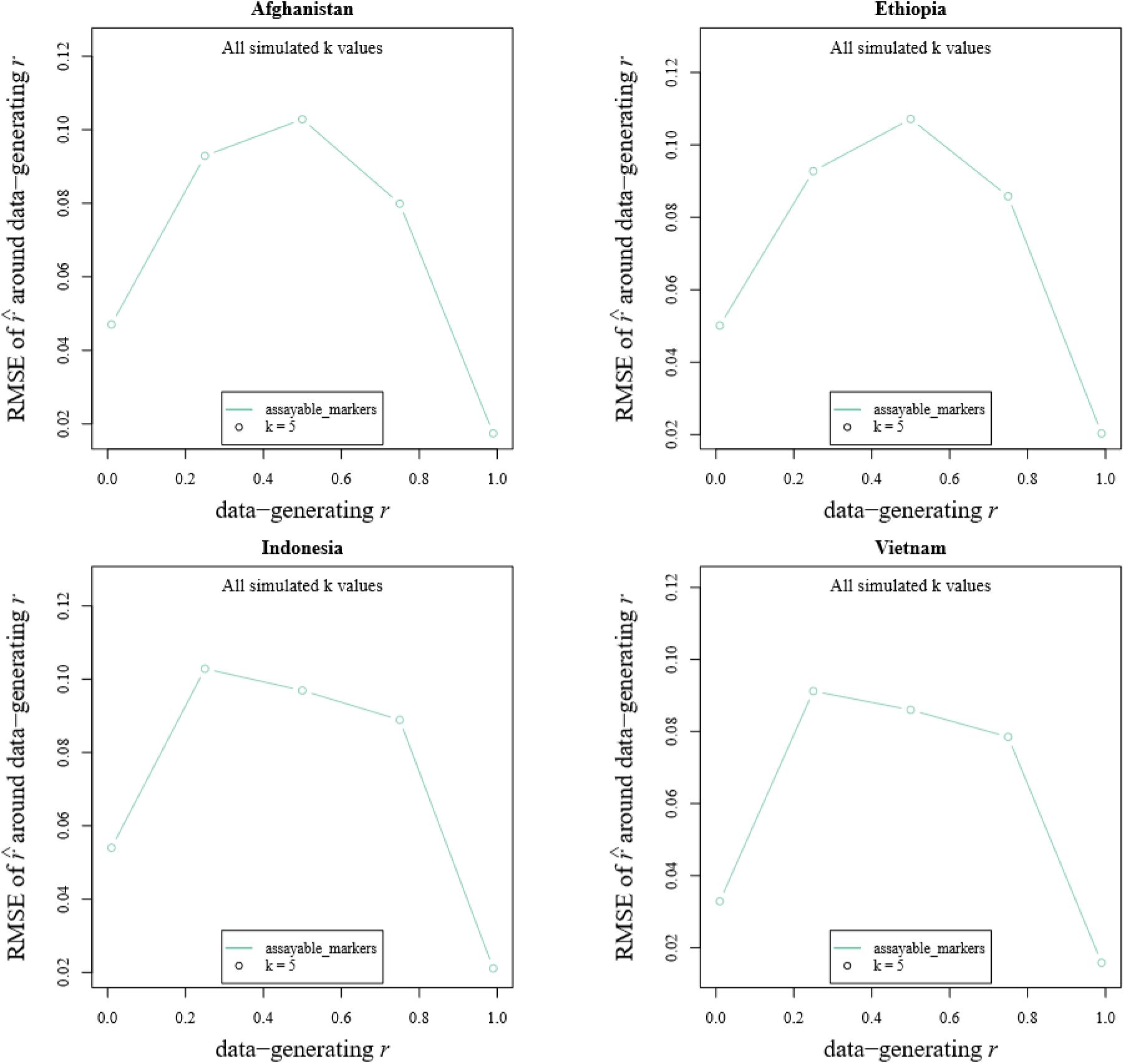
Simulations of IBD estimation in different geographic areas using the microhaplotype panel. Root mean square error (RMSE) of relatedness estimates based on data simulated using five different data-generating relatedness estimates, *r* (specifically IBD of 0 [unrelated], 0.25 [half-sibling], 0.5 [sibling], 0.75 [highly related], and 1 [clonally identical]) with switch rate parameter *k* set to 5. Data were generated using *paneljudge* software on 91 high performance microhaplotypes and 4 drug resistance markers in independent infections from Afghanistan (n=158), Ethiopia (n=213), Indonesia (n=38) and Vietnam (n=89). In all populations, half-siblings and siblings had the highest RMSE, but this remained below 0.12 in all cases.

### Potential of microhaplotype-based IBD to inform on recurrence

To demonstrate the potential of the assay to inform on the origin of recurrences, the microhaplotype-based IBD was determined using *DCifer* on data from 108 pairs of initial and recurrent infections (Supplementary Data 6). These isolates came from individuals enrolled into a randomized controlled trial at two sites in Ethiopia and treated with either chloroquine (CQ), CQ plus primaquine, artemether-lumefantrine (AL) or AL plus primaquine^16^. As illustrated in Figure 4a, the patients treated without primaquine had a higher frequency of highly related pairs (arbitrary IBD threshold ≥0.25), compared to those treated with primaquine, consistent with a greater risk of relapsing and recrudescent infections. In total, 84% (74/88) of paired isolates in patients not treated with primaquine had IBD ≥25% compared to 60% (12/20) of those treated with primaquine (*χ*2 = 4.44, *p*=0.0351). Overall, 20% (18/70) of paired isolates recurring within 120 days of treatment had low relatedness (IBD <25%) compared to 55% (11/20) recurring after 120 days; (*χ*2 = 8.22, *p*=0.004), consistent with a rising proportion of reinfections at later time points (Figure 4b).

**Figure 4.**
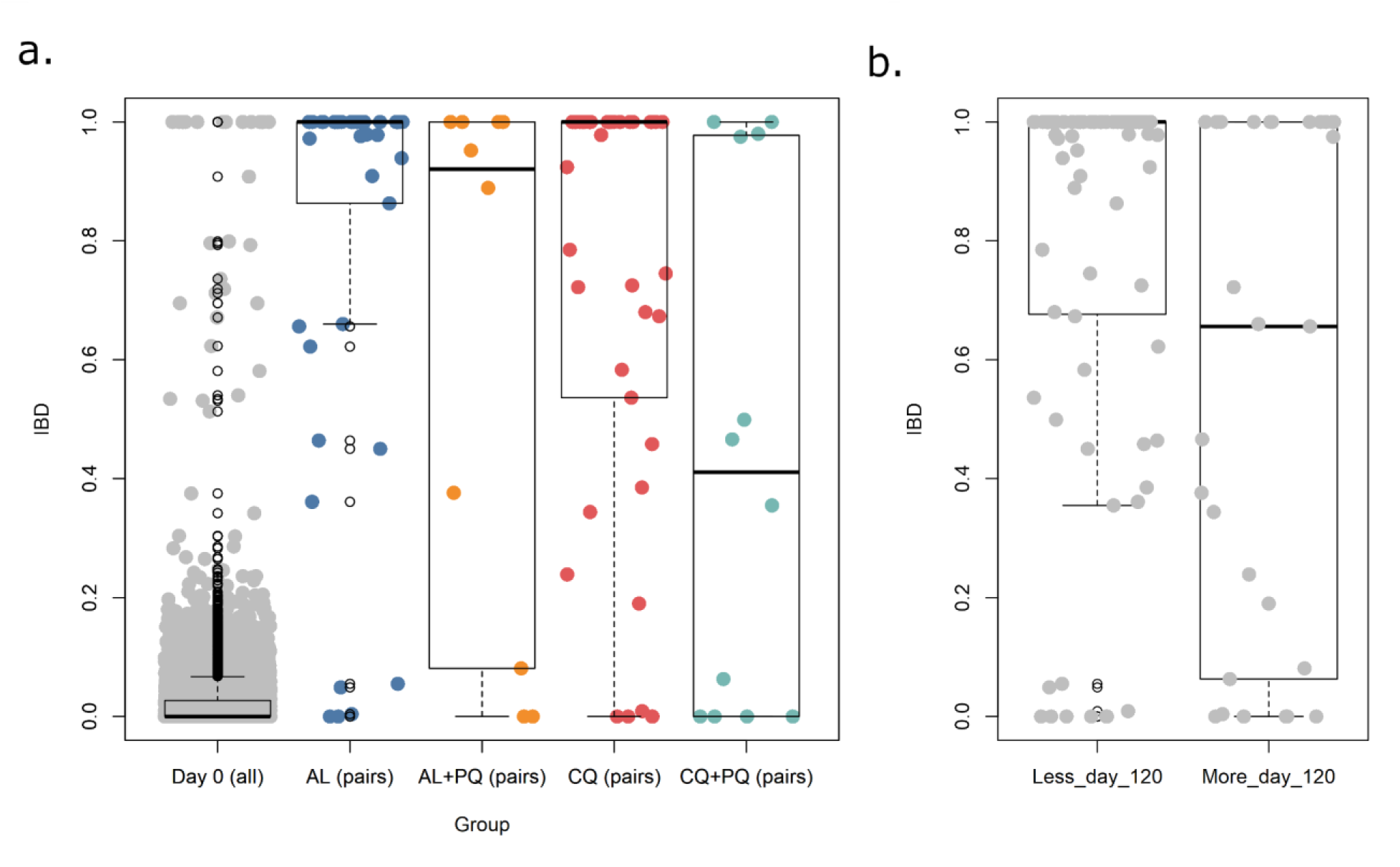
IBD distributions by treatment and time to recurrence in a randomized controlled trial conducted in Ethiopia. Panel a) presents the IBD distributions in initial and recurrent infection pairs across all pairs and grouped by treatment arm; AL (Artemether-Lumefantrine), CQ (Chloroquine), AL+PQ (AL + Primaquine) and CQ+PQ. Panel b) presents the same IBD data grouped by recurrences occurring less versus more than 120 days after the initial infection. Each boxplot presents the median, interquartile range and min and max value. Data are presented on recurrence pairs from 108 independent patients, with only one infection pair presented per patient to avoid potential bias (n=108 patients, 216 samples). Majority (86%, 93/108) of pairs reflect day 0 and recurrence 1 time points. Where the day 0 or recurrence 1 infections failed genotyping or had inconclusive clinical metadata, consecutive pairs of recurrence 2 to 4 pairs were used instead as the patients received the same treatment up to recurrence 4.

### Spatial transmission potential

The utility of the marker panel to define spatial transmission dynamics was explored using the isolates from Afghanistan, Ethiopia, Indonesia and Vietnam. As illustrated in the PCoA and neighbour-joining trees in Figures 5a and 5b, SNP-based identity-by-state (IBS) analyses on 280 infections (with <50% probability of polyclonality) demonstrated differentiation between borders. In accordance with the low eMOI distribution, isolates from Sumatra, Indonesia, revealed clusters of genetically identical infections indicative of clonal expansion. IBD analyses on the microhaplotype panel conducted in both clonal and polyclonal infections (n=498) from the same sites revealed evidence of local and inter-site transmission networks and confirmed the clonal clustering in Sumatra (Figures 6a-d).

**Figure 5.**
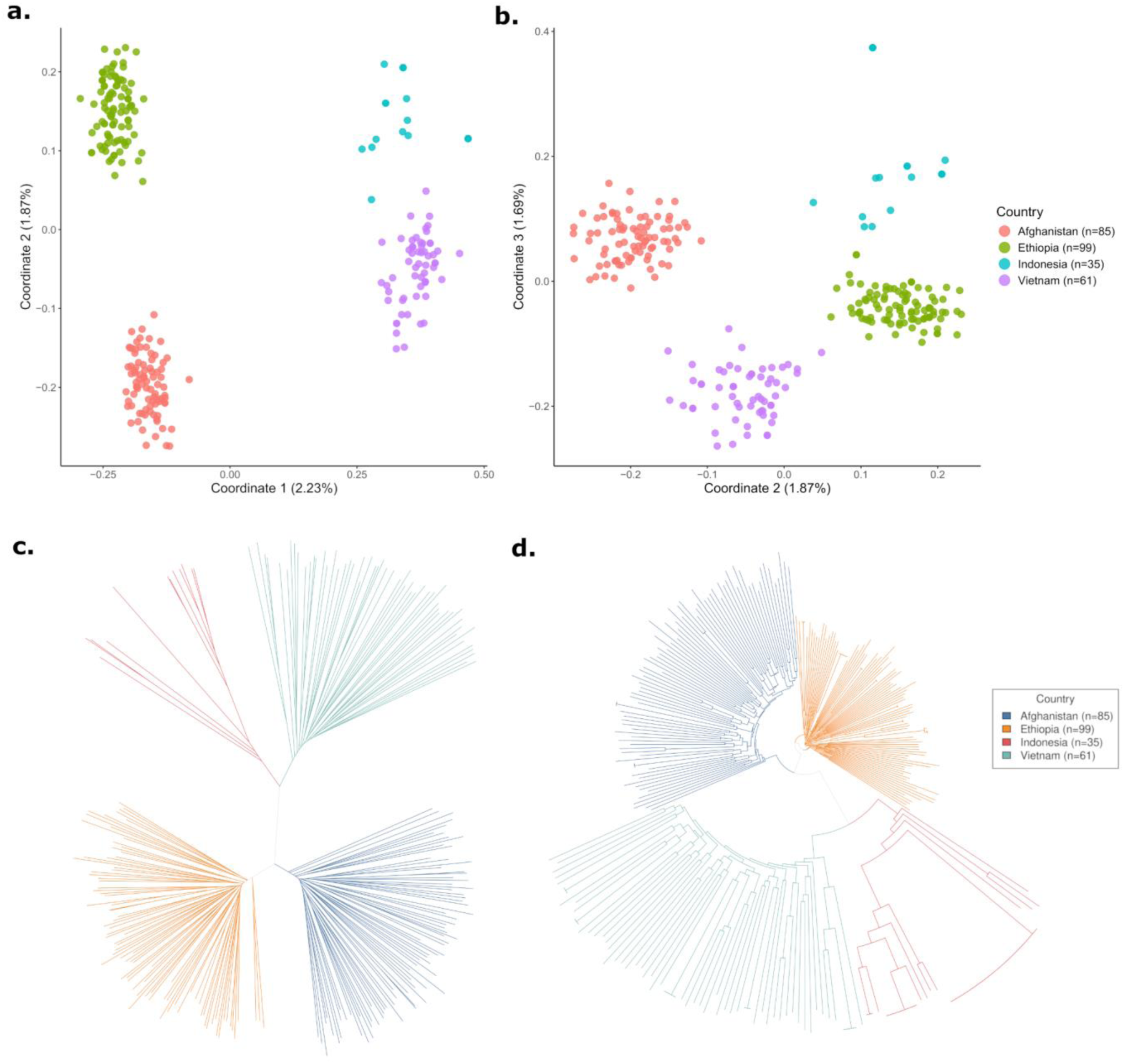
Identity-by-state-based spatial patterns. Panels a) and b) present Principal Coordinate (PCoA) plots derived from distance matrices on the microhaplotype calls. High separation of countries is observed with principal coordinates 1 and 2, with even further separation of Indonesia from Vietnam on principal coordinate 3. Panels c) and d) present unrooted and rooted, respectively, neighbour-joining trees based on the same distance matrix as per the PCoA plots. The neighbour-joining trees further illustrate distinct clustering by country and, additionally, highlight a clonally identical cluster of infections in Sumatra, Indonesia. All plots were generated using data on 280 independent, monoclonal infections.

**Figure 6.**
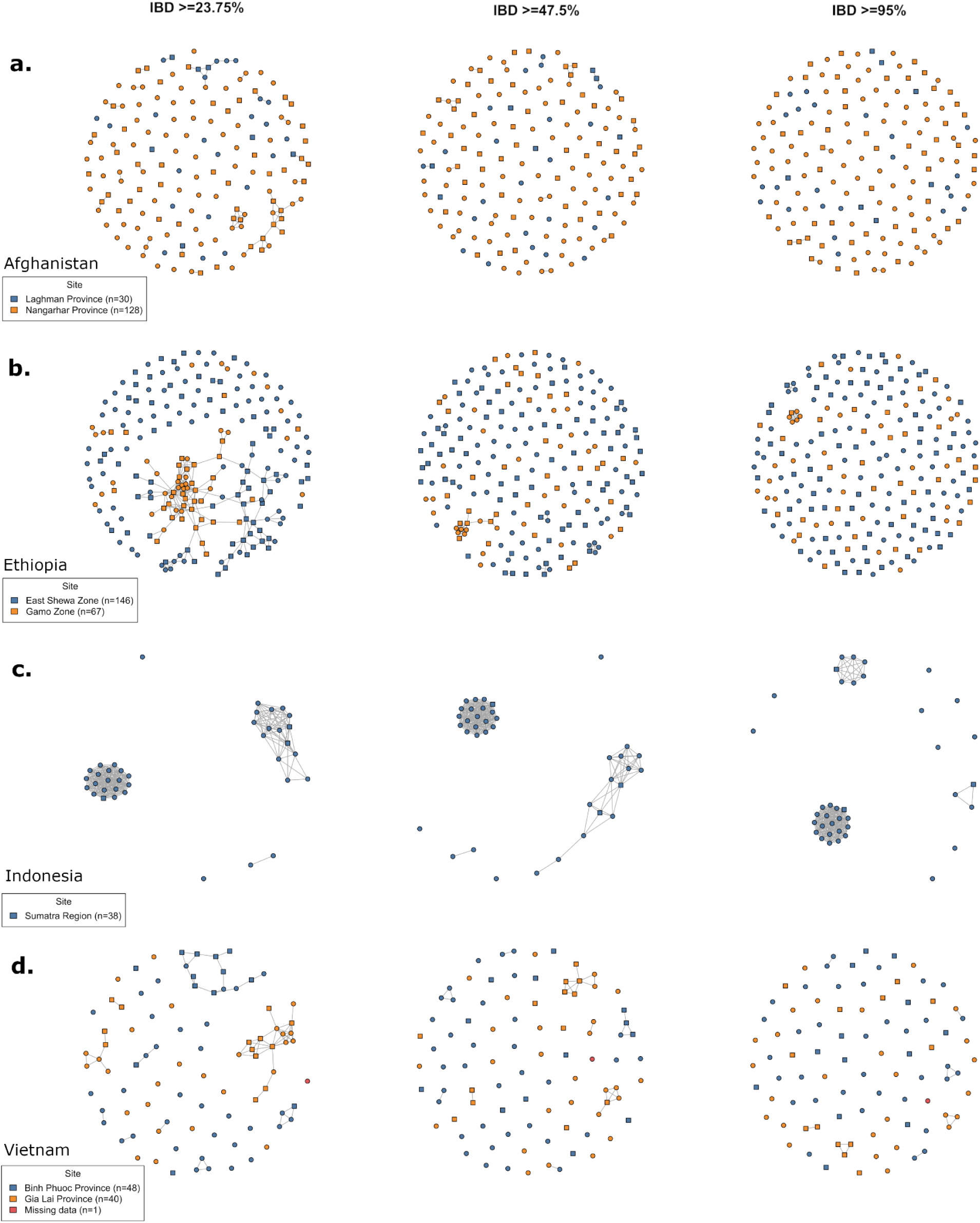
IBD-based spatial patterns using microhaplotype data. Panels a) to d) present networks illustrating IBD-based connectivity between infections in Afghanistan (a), Ethiopia (b), Vietnam (c), and Sumatra, Indonesia (d). Each shape reflects an infection, colour-coded by site, and with shapes reflecting monoclonal (circle) versus polyclonal (square) infections. For each country, connectivity (illustrated by connecting lines between shapes) is presented at IBD thresholds of minimum ∼25%, 50% and 95% (left to right). IBD measures were calculated on the microhaplotype calls using *DCifer* software. At the ∼25% IBD threshold, large networks (10 or more connected infections) are observed amongst infections from the same site, with some connectivity between networks from different sites; these networks shrink with increasing IBD in all sites except for Sumatra, Indonesia, where the networks appear to reflect clonal clusters (retained at IBD ≥95%). All plots were generated using data on independent infections: Afghanistan (n=158), Ethiopia (n=213), Vietnam (n=89) and Indonesia (n=38).

### Effective Plasmodium species confirmation

In addition to the microhaplotypes, a previously described mitochondrial amplicon was included in the assay to confirm *Plasmodium* spp. ^17^. The assay amplifies coordinates PvP01_MIT_V2:2904-3149, which include species-specific SNPs and indels. Using *P. vivax* samples from a range of countries, and *P. falciparum*, *P. malariae*, *P. ovale* spp. and *P. knowlesi* negative controls, we confirmed amplification of the mitochondrial marker at moderate to high depth and coverage in all *Plasmodium* species (range 28-5212) (Supplementary Table 4). Concordance between PCR-based and mitochondrial species classification was confirmed for each of the *Plasmodium* samples.

### Drug resistance candidates

Our assay included a non-exhaustive selection of amplicons encompassing markers of antimalarial drug resistance including the multidrug resistance 1 (*pvmdr1)* 976 and 1076 loci, and dihydropteroate synthase (*pvdhps* ) 383 and 553 loci that have been associated with *ex vivo* or clinical phenotypes (see ^18^). The dihydrofolate reductase (*pvdhfr*) 57, 58, 61 and 117 loci did not multiplex effectively with the other markers based on bioinformatic predictions and were thus not included. The prevalence of the variants by population is summarized in Figure 7a and Supplementary Table 5. The prevalence of the *pvmdr1* Y976F variant, the most widely characterized candidate of CQR, ranged from 0% in Afghanistan to 100% in Sumatra, Indonesia ^19^. The F1076L variant, which has also been implicated in CQR, exceeded 90% frequency in all 4 countries. The prevalence of A383G mutation in *pvdhps*, a marker of antifolate resistance, varied highly, ranging from 2% in Afghanistan to 80% in Vietnam. No *pvdhps* A553G mutations were observed in any country.

**Figure 7.**
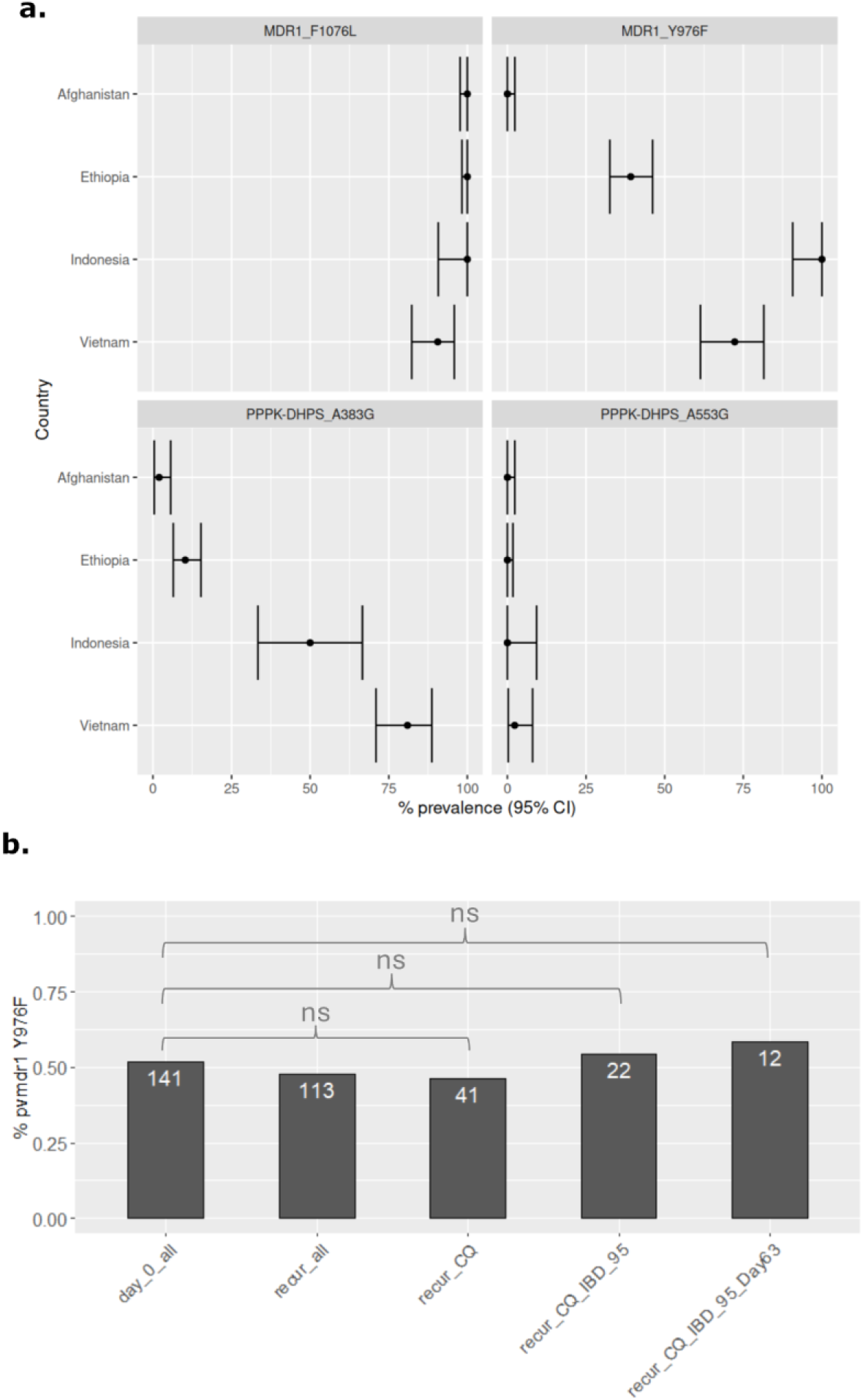
Amino acid frequencies at *P. vivax* drug resistance candidates. Panel a) presents proportions and corresponding 95% confidence intervals (CIs) for the given amino acid changes in baseline population samples from each country. All frequencies reflect the suspected drug-resistance-conferring amino acid. All plots were generated using independent samples (n=498). Panel b) presents the proportions of the *pvmdr1* Y976F variant, which has been associated with CQ resistance, in the respective patient groups within the Ethiopian randomized control trial; all genotyped baseline (day 0) infections irrespective of treatment arm (day_0_all), all recurrent infections irrespective of treatment arm (recur_all), all recurrent infections in the CQ only arm (recur_CQ), all recurrent infections in the CQ only arm with IBD>0.95 relative to the prior infection (enhancing probability of recrudescence based on genetics; recur_CQ_IBD_095), and all recurrent infections in the CQ only arm occurring prior to day 63 and with IBD>0.95 relative to the prior infection (enhancing probability of recrudescence based on genetic and time-to-event metadata, recur_CQ_IBD_095_Day63). Sample sizes are annotated in white font over the respective bars. ns = non-significant difference in proportion between the indicated groups.

We also explored the prevalence of the *pvmdr1* Y976F variant in the Ethiopian RCT cases (Supplementary Data 6, Figure 7b). Amongst 141 day 0 samples with successful microhaplotype and *pvmdr1* 976 genotyping, Y976F prevalence was 52% (73/141). The prevalence of Y976F mutants amongst recurrences occurring before day 63 in the CQ only arm with IBD >=0.95 relative to the prior infection (likely CQ recrudescence) was not significantly different from the day 0 samples, but sample size was small (7/12, 58%, chi-square, *p*=0.892).

## Discussion

We have established a highly multiplexed rhAmpSeq assay (up to 98 amplicons) to address critical knowledge gaps in our understanding of antimalarial efficacy and transmission dynamics of *P. vivax*. Our assay can be applied to low-density infections and to NGS platforms that are generally available in reference laboratories in malaria-endemic settings.

A critical requirement of the assay was its ability to be implemented in surveillance frameworks in vivax-endemic countries. This necessitated high sensitivity to genotype low-density *P. vivax* infections at affordable cost using locally accessible sequencing platforms. With full chemistry, our analyses of serial dilutions demonstrated yields of >100 mean read depth when parasite density was >70 per microliter whole blood. Defining a meaningful assay sensitivity with amplicon sequencing approaches is challenging as several factors other than parasite density can impact on read depth, including the yield of the sequencing platform used, the level of sample multiplexing, the relative target biomass of other samples on the same run, and whether a pre-amplification step is applied. To stress-test our assay, we evaluated the sensitivity of target detection under conditions at the lower end of potential sequence yield at the benefit of lower processing cost. Sensitivity evaluations were undertaken on a MiSeq platform, which is the most widely available platform in malaria endemic settings. The paired end read yield per run is approximately 20-30 million for the MiSeq compared to 260-800 million and up to 40 billion on the NextSeq and NovaSeq platforms, opening the possibility for even greater yield if the setting permits. Furthermore, samples could be pooled and did not require pre-amplification with selective Whole Genome Amplification (sWGA) or Primer Extension Preamplification (PEP) ^20,21^. Our rationale was that the rhAmpSeq RNase H2 enzyme-dependent amplicon sequencing chemistry should reduce the need for pre-amplification steps relative to standard AmpSeq as it minimizes spurious primer-primer interactions and off-target amplification ^22^.

Another essential data analysis requirement of our assay was the ability to generate genetic data that effectively capture pairwise IBD in *P. vivax* infections with the aim of defining the origin of recurrent infections. Previously described *P. vivax* amplicon sequencing assays, such as a widely used 42-SNP barcode, have not comprised the density of SNPs needed to capture IBD accurately ^8,23,24^. Previously using *in-silico* methods, we demonstrated that panels of ∼100 *P. vivax* microhaplotypes can capture pairwise IBD states ranging from 0 (unrelated) to 1 (clonally identical) with low error (RMSE under 0.12)^8^. In our current study, we applied the same *in-silico* models to demonstrate that, despite less optimal spacing and slightly fewer markers than in the *in-silico* panels, our assayable set of microhaplotypes has comparably low error in IBD estimation at simulated states of 0, 0.25, 0.5, 0.75 and 1 (RMSE under 0.12 with 92 microhaplotypes). We also note the adaptability of the rhAmpSeq protocol, enabling addition of new markers, which could be selected to cover genomic regions with low marker density. The software developed in our previous study supports microhaplotype selection that can be customized to specific populations or for specific marker traits ^8^.

In addition to the *in-silico* analysis, we explored microhaplotype-based IBD patterns between pre and post treatment peripheral blood isolates collected from a clinical trial conducted in Ethiopia ^16^. Our results demonstrated a higher proportion of highly related but non identical (IBD 0.25-0.95) paired isolates in patients not treated with primaquine, which we postulate reflects a greater risk of relapses in the absence of radical cure. We also observed a higher frequency of clonally identical infection pairs (IBD ≥0.95) in patients not treated with primaquine, likely reflecting a combination of relapses and recrudescence events, that can be further dissected using time-to-event analyses ^25^. Lastly, we observed a greater frequency of paired infections with low relatedness (IBD <0.25) after 4 months consistent with an increasing risk of reinfections during prolonged follow-up after antimalarial treatment. These findings concur with expectations and thus contribute proof of concept on the utility of genetics to inform on recurrence. Data from known relapse events, i.e. from individuals who were relocated to non-endemic areas, will be needed to confirm these findings. Further work is also needed to dissect the potential contribution of the splenic reservoir to recurrent *P. vivax* infections. Lastly, the incorporation of genetic data with time-to-event information using modeling approaches that can handle polyclonal infections, such as the PV3Rs software will greatly inform recurrence classification (https://github.com/aimeertaylor/Pv3Rs). The data have been made open access to support further developments in this space.

The microhaplotype data produced in this study also provides evidence of the potential for our assay to inform spatial patterns of diversity and connectivity. At a macro-epidemiological level, there was a distinct separation of populations by national boundaries, that could facilitate detection of cross-border importation of infection. Within countries, trends in within-host diversity and connectivity highlighted different transmission dynamics in Sumatra, Indonesia, relative to other sites. Relatively low prevalence of polyclonal infections and large clonal clusters supported low transmission and subsequent inbreeding in Sumatra relative to the other sites. The observed patterns infer that Sumatra is approaching the pre-elimination phase and may be receptive to targeted rather than broad scale intervention approaches. Within countries, we also observed evidence of moderate to large local transmission networks, largely confined to specific sites but with some connections across sites. As the data repositories grow, it should be possible to better define the major transmission networks within local regions and their epidemiological context to inform on major reservoirs of infection and routes of infection spread between communities.

Our panel included a non-exhaustive selection of *P. vivax* drug resistance candidates. The markers and mechanisms of resistance to the more widely used drugs such as chloroquine, artemisinin and partner drugs are not well understood in *P. vivax* ^5^. The inference on treatment failure that can be made on the available markers, including those incorporated in our panel, is therefore limited from an NMCP perspective. However, the adaptability of the rhAmpSeq protocol ensures that new drug resistance markers can be readily added to the panel as they arise. Insights from the current selection of candidates included notable variation between countries in the prevalence of the *pvmdr1* Y976F variant, which has been implicated as a minor modulator of chloroquine (CQ) resistance ^19^. The Y976F variant was present at 100% frequency in Sumatra, Indonesia. To our knowledge, this is the first report on Y976F in Sumatra, and aligns with the high prevalence (100%) of this variant in Papua province, Indonesia, a region that has historically harbored high-grade CQ resistant *P. vivax* infections ^19,26,27^. Although not at fixation, high prevalence of the Y976F variant was also observed in Vietnam (73%) and Ethiopia (41%), but not in Afghanistan (0%). Therapeutic efficacy surveys report low level (<10%) CQ resistance in Ethiopia, Vietnam and Afghanistan, while 16-65% failures by day 28 have been reported in Sumatra, Indonesia ^28–36^. The connection between the Y976F variant and clinical CQ efficacy is also unclear. Our exploration of the Y976F variant in recurrent cases from the CQ only arm of an RCT conducted in Ethiopia in 2015 revealed a non-significant difference in prevalence (58%) relative to the baseline study set (51%). However, the sample size was small, with only 12 recurrence pairs in the CQ arm meeting the IBD and pre-day 63 requirement. Further exploration of phenotypic associations utilizing IBD and time-to-event data are needed in endemic settings with evidence of higher grade CQ resistance.

The panel also includes a previously described mitochondrial amplicon, comprising SNPs and indels that support *Plasmodium* spp. determination ^17^. We used the amplicon to successfully confirm *Plasmodium* spp. in a selection of non-*P. vivax* controls. A caveat of the assay is the inability to distinguish *P. ovale curtisi* from *P. ovale wallikeri*. However, this could be addressed with the addition of other mitochondrial amplicons ^37^. Although most assays displayed high specificity to *P. vivax*, several primers yielded amplicons of *P. knowlesi* DNA with high coverage (>0.9) and moderate to high read depth.

Although *P. knowlesi* is less common in Southeast Asia outside of Malaysian and Indonesian Borneo and considering that *P. vivax* has been eliminated from Malaysia, the *Plasmodium* spp marker may still be helpful to detect potential mixed *P. vivax* and *P. knowlesi* infections which could yield inaccurate genotypes at the specified markers.

Library preparation costs were approximately $AUD 46 per sample for our study but can be approximately halved (to $AUD 23) by using half-chemistry reaction volumes for PCR 1. We observed similar sensitivity between the full- and half-chemistry reactions in our serial dilution experiments.

In summary, the tools generated in our study provide a major in-road to establishing high-throughput genetic data on *P. vivax* at reasonable cost that can provide policy relevant information on transmission dynamics, parasite reservoirs and the spread of infection across national and provincial borders.

## Methods

### Marker selection and assay design

193 microhaplotype markers, 5 drug resistance candidates, and two mitochondrial *Plasmodium* species-confirmation markers, were selected for assay design (Supplementary Data 1). The set of 193 microhaplotypes were derived from two 100-microhaplotype panels (referred to as the high-diversity and the random microhaplotype panels respectively based on the SNP filtering methods) that we previously demonstrated exhibit high accuracy in IBD determination ^8^. The rationale for selecting two panels was based on expectation that some markers in the preferred panel (the high-diversity panel) might not be assayable. The drug resistance candidates were not chosen to reflect an exhaustive list of resistance candidates but rather a selection of SNPs that have previously been associated with *ex vivo* or clinical phenotypes ^18^. The species-confirmation markers have been described in a previous malaria amplicon panel ^17^. Primers and multiplex pools were designed by Integrated DNA Technologies (IDT). Primer specifications included non-mapping to the human (GRCh38.p14), *P. falciparum* (Pf3D7), *P. malariae* (PmUG01), *P. ovale wallikeri*/*curtisi* (GH01), and *P. knowlesi* (PKNH) reference genomes. A variant calling format (VCF) file for the open-access MalariaGEN Pv4.0 dataset, comprising 911,901 high-quality variants at 1,895 global *P. vivax* samples, was also provided to confirm non-mapping to highly variable regions of the *P. vivax* genome ^38^. A set of 148 markers that met the described primer specifications, and compatible within a single-plex reaction, were taken forward for a pilot experimental run, comprising 176 positive and 16 negative controls (Supplementary Data 1). From the pilot data (not presented here), a set of 98 markers were selected for the final assay (Supplementary Data 1). Requirements for the final panel included high specificity to *P. vivax* and approximate uniform microhaplotype distribution across the genome (Supplementary Figure 1).

### Patient samples

The sensitivity and specificity of the 98-plex assay was evaluated using *P. vivax*-infected serial blood dilutions and unmodified *P. vivax*-infected blood samples, stored as dried blood spots or refrigerated whole blood in EDTA-coated microtainer or vacutainer tubes. The dilution series were prepared by mixing each of the three *P. vivax*-infected whole blood samples (microscopy-determined parasite densities of 740 to 960 parasites/ul blood) with uninfected human whole blood at 10-fold serial dilutions (0.7 to 0.96 parasites/ul blood). Whole blood samples from uninfected human (n=3), *P. falciparum* (n=2), *P. malariae* (n=2), *P. ovale* (n=1), and *P. knowlesi* (n=3) were included as negative controls. The samples were collected within the framework of previously described clinical trials and cross-sectional surveys, as well as returning travelers presenting with malaria at the Royal Darwin Hospital, Australia, and represented 8 vivax-endemic countries (Supplementary Data 2) ^39–42^. DNA extraction was undertaken using Qiagen’s QIAamp kits for respective dried blood spots or whole blood.

### Real-time PCR

The cycle threshold (Ct) values of *P. vivax* DNA samples were assessed using an Applied Biosystems® QuantStudio™ 6 Flex Real-Time PCR System (ThermoFisher Scientific) with a TaqMan real-time PCR-based assay, as adapted from ^43^. This assay detects *P. vivax* by targeting a conserved region of the mitochondrial cytochrome oxidase 1 gene (*pvmtcox1*) ^43^. Each qPCR reaction mixture consisted of 10 ul TaqMan Universal Mix II master mix (ThermoFisher Scientific), 1.6 ul of both the forward and reverse *Pvmtcox1* primers (10 uM), 0.8 µL of the *pvmtcox1* probe (10 uM), 2 ul H2O, and 4 ul gDNA. The thermocycling conditions included an initial denaturation at 95°C for 10 min, followed by 45 cycles of denaturation at 95°C for 15 sec, and annealing and elongation at 60°C for 1 min. Data analysis was performed using QuantStudio RT-PCR software to derive the threshold cycle (Ct) values.

### Whole genome sequencing (WGS) data

A combination of newly generated and pre-existing WGS data was used in the study (Supplementary Data 7). The pre-existing WGS data were derived from the MalariaGEN Pv4.0 repository ^38^. Briefly, the data were generated using 100-150 bp paired-end sequencing on a HiSeq or MiSeq instrument (Illumina). The resultant reads were analyzed with the biallelic SNP pipeline used in this study, which incorporated read mapping against the *P. vivax* P01 reference using BWA-MEM2 and variant calling using the Genome Analysis Toolkit v4.5 ^44,45^. A total of 911,901 high-quality biallelic SNP loci were derived. Positions with less than 5 reads were considered genotype failures, with a minimum of two reference and two alternate alleles, and a minimum of 10% of minor allele read frequency required to define a genotype as heterozygous. The new WGS data were generated using microscopy-positive *P. vivax* infections collected within a therapeutic efficacy survey of chloroquine, conducted in Arbaminch, Ethiopia in 2019. Briefly, DNA was extracted from 1-2 ml white blood cell-depleted (Plasmodipur-filtered) whole blood samples using Qiagen’s QIAamp blood midi kits. Library preparation and sequencing was conducted on the gDNA at the Wellcome Sanger Institute using the Illumina HiSeq platform, generating 150bp paired reads. The resultant reads were mapped against the PvP01 reference genome and genotype calling was conducted using the 911,901 MalariaGEN Pv4.0 SNPs, following the same methods. Only samples with more than 50% genotype calls were included in the analysis.

### rhAmpSeq library preparation and sequencing

Library preparation of the 98-plex marker panel was conducted using IDT’s rhAmpSeq methodology. Details on the protocol are provided in Supplementary Note 1 and on the protocols.io website (https://www.protocols.io/file/rfffce597.pdf). In brief, genomic DNA samples were subject to a single multiplexed (all 98 targets amplified within one pool) primer amplification using pre-balanced, customized, rhAmpSeq primers. The product was diluted and subject to a second PCR, incorporating indexes and Illumina sequencing adaptors. The products from the second PCR step were pooled (incorporating up to 384 samples), resulting in the final library. The library was bead purified, assessed for quality (fragment size and quantity), and sequenced with 150 bp paired-end clusters on a MiSeq instrument (Illumina).

### Amplicon data read mapping and variant calling

Options for variant calling from fastq files at both biallelic SNPs and multiallelic microhaplotypes were incorporated into the data processing pipeline, which is available at https://github.com/vivaxgen/MicroHaps and described in Supplementary Note 2. In short, for the biallelic SNP pipeline, vcfs are generated using a pipeline that conducts read mapping via *bwa-mem2*, and variant calling with GATK v4.5. Positions with less than 25 reads were considered genotype failures, with a minimum of two reference and two alternate alleles, and a minimum of 10% of minor allele read frequency required to define a genotype as heterozygous. For the microhaplotype-calling pipeline, we adapted an existing *P. falciparum* microhaplotype pipeline which utilizes the *DADA2* software for denoising ^46^. The microhaplotype pipeline includes adapter and primer removal with *cutadapt*, and custom scripts to postprocess *DADA2* output.

### Population genetic measures

For the resulting microhaplotype data, within-host infection diversity was characterized by the effective multiplicity of infection (eMOI) metric using *MOIRE* software ^47^. For the WGS data, within-host diversity was characterized using the *F*_WS_ score, calculated from biallelic SNP loci ^48^. The R-based *paneljudge* package was used to assess the relative mean square error (RMSE) of the microhaplotype markers at IBD levels of 0 (unrelated), 0.25 (half-siblings), 0.5 (siblings), 0.75 and 1.0 (identical) (https://github.com/aimeertaylor/paneljudge). Between-sample IBD was measured using *DCifer* ^49^. Network plots were generated using the *R*-based *igraph* package (https://github.com/igraph). Identity-by-state (IBS)-based measures of genetic distance were calculated using the *R*-based ape package, treating the microhaplotypes as multiallelic variants, and presented as neighbour-joining trees and principal coordinate (PCoA) plots ^50^.

### Plasmodium speciation based on genetic distance matrix

To confirm the *Plasmodium* spp., amplicons targeting a universal mitochondrial sequence region were mapped to a synthetic genome. The synthetic genome was constructed using reference genomes of five *Plasmodium* species: PvP01_MIT_v2:2904-3149 (*P. vivax*), PmUG01_MIT_v1:1115-1361 (*P. malariae*), PKNH_MIT_v2:1632-1875 (*P. knowlesi*), Pf3D7_MIT_v3:1634-1877 (*P. falciparum*), and PocGH01_MIT_v2:2894-3137 (*P. ovale*). The synthetic genome was generated through an alignment containing the specified regions within the reference genomes; aligned using muscle ^51^. For each sample with more than 10 reads and >0.9 coverage in the mitochondrial sequence region, the reads were mapped to the synthetic genome. The mapped reads were used to call a consensus sequence using iVar ^52^. The consensus sequence was then compared pairwise to each sequence within the multi-sequence alignment, containing all the reference genomes. The species of the sample was determined based on the highest pairwise alignment score.

### Inclusion and ethics

The authors in this study combine researchers from the malaria-endemic countries represented in the study (Afghanistan, Bangladesh, Colombia, Ethiopia, Indonesia and Vietnam) and nonmalaria endemic areas. The researchers from malaria-endemic countries were involved throughout the research process to ensure that the assays are implementable and impactful in local communities affected by malaria. All samples in this study were derived from blood samples obtained from patients positive for malaria and collected with informed consent from the patient, or patient parent / legal guardian where individuals were less than 18 years of age. At each location, sample collection was approved by the appropriate local institutional ethics committees. The following committees gave ethical approval for the partner studies: Human Research Ethics Committee of NT Department of Health and Families and Menzies School of Health Research, Darwin, Australia; Islamic Republic of Afghanistan Ministry of Public Health Institutional Review Board, Afghanistan; ICDDR,B Ethical Review Committee, Bhutan; Research Ethics Board of Health, at the Ministry of Health in Bhutan; Comite Instiucional de Etica de Investigaciones en Humanos, Colombia; Comite de Bioetica Instituto de Investigaciones Medicas Facultad de Medicina Universidad de Antioquia, Colombia; Armauer Hansen Research Institute Institutional Review Board, Ethiopia; Addis Ababa University College of Natural Sciences, Ethiopia; Addis Ababa University, Aklilu Lemma Institute of Pathobiology Institutional Review Board, Ethiopia; National Research Ethics Review Committee of Ethiopia; Eijkman Institute Research Ethics Committee, Jakarta, Indonesia; Research Review Committee of the Institute for Medical Research and the Medical Research Ethics Committee (MREC), Ministry of Health, Malaysia; Scientific and Ethical Committee of the Hospital for Tropical Diseases in Ho Chi Minh City, Vietnam; The Ministry of Health Evaluation Committee on Ethics in Biomedical Research, Vietnam.

## Data Availability

The MalariaGEN Pv4.0 WGS data are available through the European Nucleotide Archive. The WGS reads from the newly generated, high-quality, Ethiopian P. vivax infections are available through the European Nucleotide Archive. The ENA accession numbers for all WGS samples used here are listed in Supplementary Data 7. All high-quality (pass) microhaplotype-based parasite reads will be made available in the European Nucleotide Archive at publication (Supplementary Data 2).

https://www.malariagen.net/data_package/open-dataset-plasmodium-vivax-v4-0/

## Data availability

The MalariaGEN Pv4.0 WGS data are available through the European Nucleotide Archive ^38^. The WGS reads from the newly generated, high-quality, Ethiopian *P. vivax* infections are available through the European Nucleotide Archive. The ENA accession numbers for all WGS samples used here are listed in Supplementary Data 7. All high-quality (pass) microhaplotype-based parasite reads will be made available in the European Nucleotide Archive at publication (Supplementary Data 2).

## Code availability

The pipelines for retrieving SNP data in variant call format (vcf), as well as microhaplotype data in Compact Idiosyncratic Gapped Alignment Report (cigar) string format, are available on GitHub at https://github.com/vivaxgen/MicroHaps.

## Acknowledgements

This work was supported, in whole or in part, by the Bill & Melinda Gates Foundation INV-043618 (supporting S.A., D.N and R.N.P). Under the grant conditions of the Foundation, a Creative Commons Attribution 4.0 Generic License has already been assigned to the Author Accepted Manuscript version that might arise from this submission. The study was also supported by the National Health and Medical Research Council of Australia (APP2001083 supporting S.A. and S.V.S.). The whole genome sequencing component of the study was supported by the Medical Research Council and UK Department for International Development (award number M006212 to D.K.) and the Wellcome Trust (award numbers 206194 and 204911 to D.K.). The OPRA clinical trial was supported by the Bill & Melinda Gates Foundation (OPP1054404 awarded to R.N.P.). We thank the patients who contributed their samples to the study, and the health workers and field teams who assisted with the sample collections. Whole genome sequencing was undertaken by the Wellcome Sanger Institute, and amplicon sequencing was undertaken at the Australian Genome Research Facility (AGRF) and Menzies School of Health Research. We thank the staff of the Wellcome Sanger Institute and AGRF for contributions to samples logistics, sequencing and informatics. We thank Ruchit Panjal for his assistance with the pipeline set up.

## Author contributions

S.A., D. N. and R.N.P. conceived the study. S.A., M.K., A Rumaseb, E.S., and H.T. designed the study. A. Rumaseb, T.P., D.H., G.W., S.V.S, N.T.T.N. and S.A. contributed to the laboratory assay design. A Rumaseb, T.P., D.H., A.O. and A Rai contributed to genotyping data generation. S.V.S, R.P., R.A. and D.P.K. contributed whole genome sequencing data production and informatic support. M.K., H.T., A.O., P.M. and D.N. contributed to bioinformatic pipeline development. M.K., E.S., H.T., K.S.H. and S.A. conducted data analysis. E.D.B., N.T.T.N, N.H.C., A.A., T.S.D., D.T.A., A.G.R., A.P.P., I.S., M.S.A., Z.P., T.L., D.E., T.W., N.A., M.G., N.D., N.J.W., D.P.K. and R.N. contributed essential field-based malaria collections and metadata, and guidance on the study design and interpretation.

