## Supplementary for "Microhaplotype deep sequencing assays to capture *Plasmodium vivax* infection lineages"

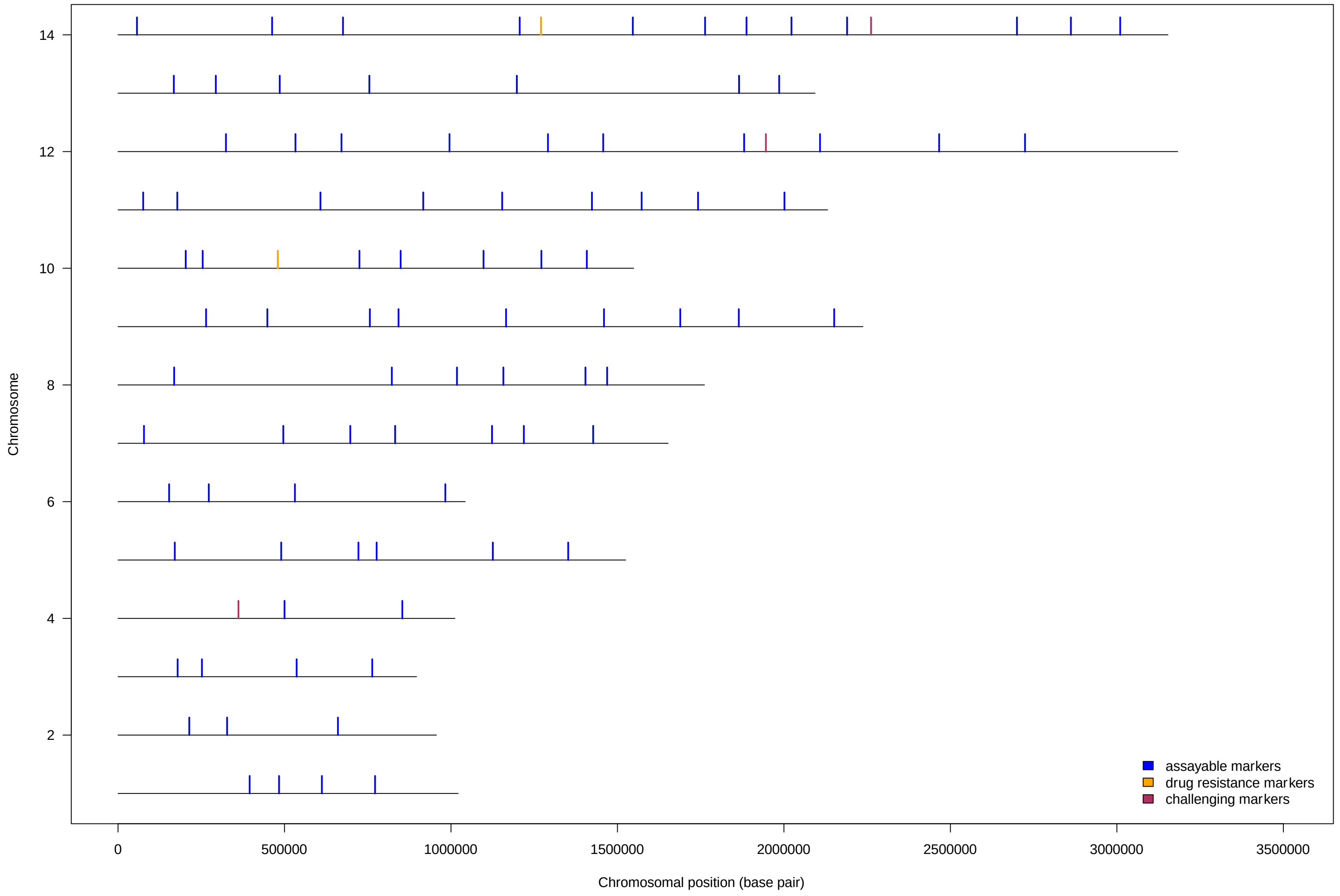

**Supplementary Figure 1. Genomic locations of the 97 nuclear genome markers.** Note, excluding the mitochondrial locus. Markers described as “challenging” displayed low read-pair depth with *dada2* output; markers 64721 (Chromosome 4), 354590 (Chromosome 12) and 466426 (Chromosome 14) (see Figure 2b).

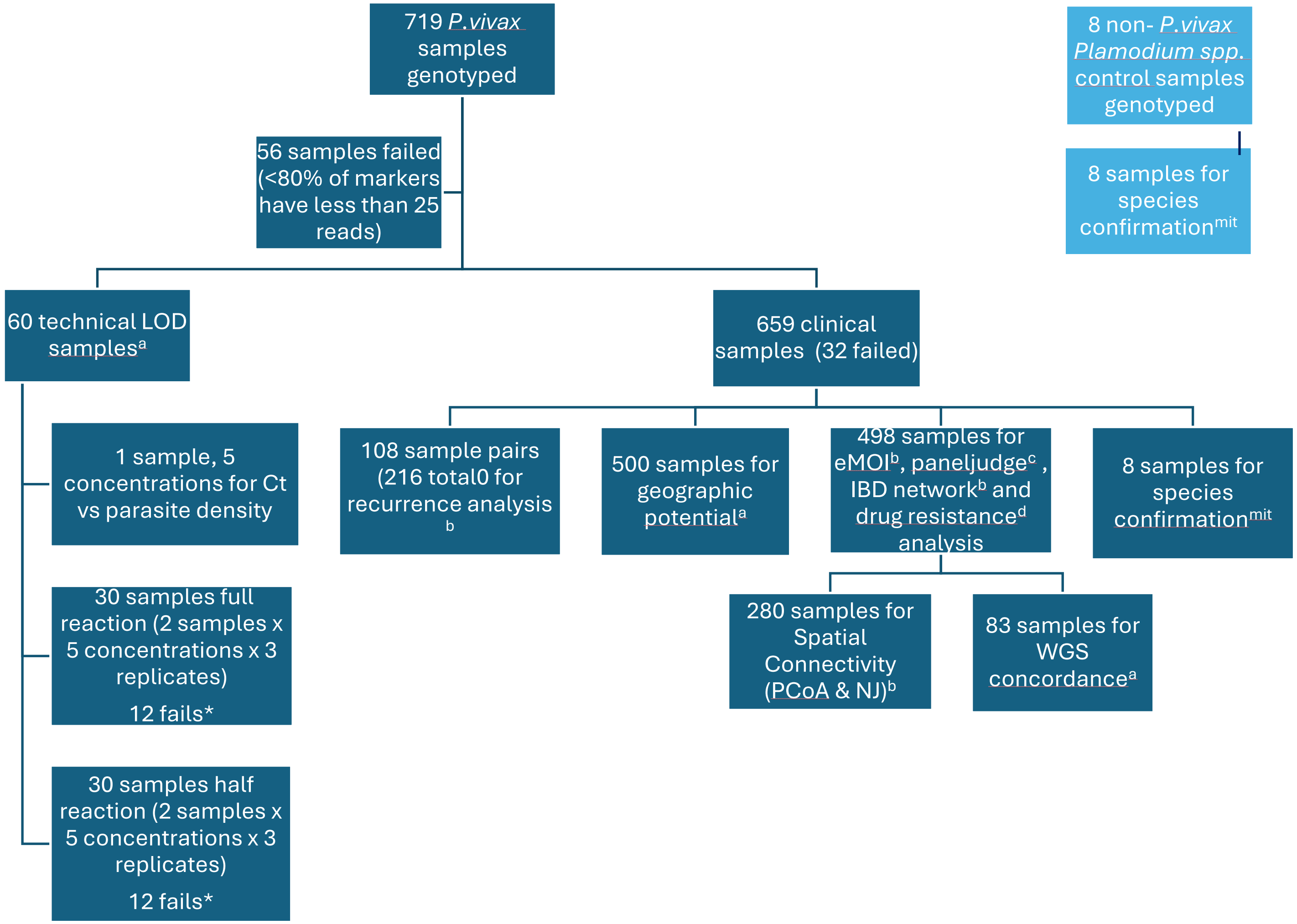

**Supplementary Figure 2. Overview flowchart of Plasmodium spp sample genotyping and analysis.** * Samples failed here were the 2 lowest concentrations (all under 9.6 ng/ul) for all three replicates. [a]97 markers used (all except mitochondria). [b] 92 markers used (excluding mitochondria, Mhap marker 354590 and 4 drug resistance candidate markers). [c] 91 markers used (excluding mitochondria, Mhap markers 354590 and 419038, and 4 drug resistance candidate markers). [d] Only markers in *pvmdr1* and *pvdhps*. [mit] only the mitochondrial region.

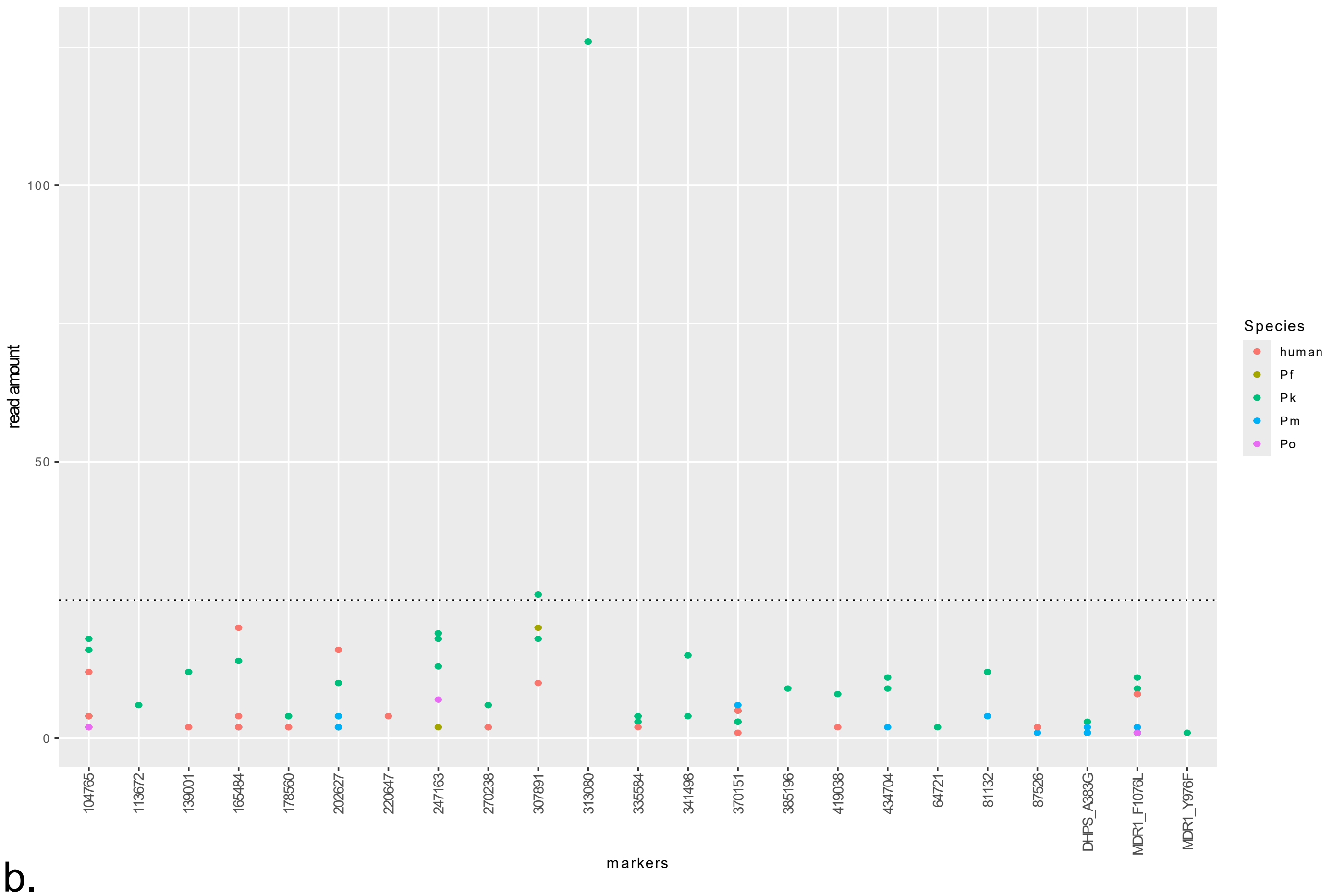
read amount
100
50
0
markers
104765
113672
139001
165484
178560
202627
220647
247163
270238
307891
313080
335584
341498
370151
385196
419038
434704
64721
81132
87526
DHPS_A383G
MDR1_F1076L
MDR1_Y976F
Species
human
Pf
Pk
Pm
Po

b.

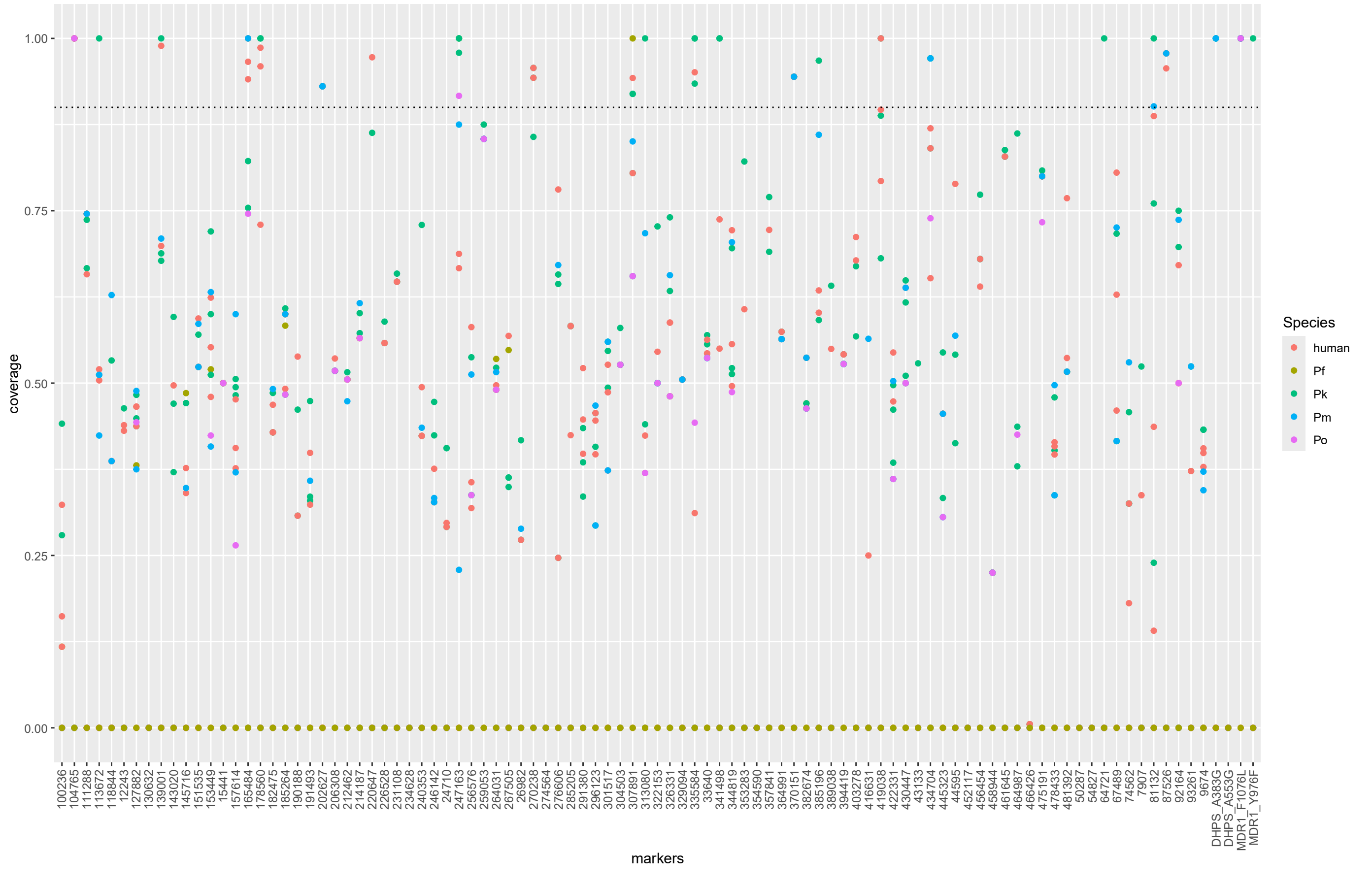
coverage
1.00
0.75
0.50
0.25
0.00
markers
100236
104765
111288
113672
118844
12243
127882
130632
139001
143020
145716
151535
153449
15441
157614
165484
178560
182475
185264
190188
191493
202627
206308
212462
214187
220647
226528
231108
234628
240353
246142
24710
247163
256576
259053
264031
267505
26982
270238
274564
276606
285205
291380
296123
301517
304503
307891
313080
322153
326331
329094
335584
33640
341498
344819
353283
354590
357841
364991
370151
382674
385196
389038
394419
403278
416631
419038
422331
430447
43133
434704
445323
44595
452117
456454
458944
461645
464987
466426
475191
478433
481392
50287
54827
64721
67489
74562
7907
81132
87526
92164
93261
9674
DHPS_A383G
DHPS_A553G
MDR1_F1076L
MDR1_Y976F
Species
human
Pf
Pk
Pm
Po

**Supplementary Figure 3. Specificity of the assays.** Panel a) presents coverage over all markers aside from the mitochondrial *Plasmodium* spp. marker in non-*P. vivax* samples. Samples with coverage over 0.9 (90%; dotted line) were further investigated for read amount in panel b), which presents the read depth of markers from non-*P. vivax* samples with coverage over 90%. Cut off (dotted line) was set to 25 for further analysis to remove background noise. The results comprise 96-plex and 384-plex runs as majority of the negative controls were processed on a pilot 96-plex run; in theory, this enhances the potential to capture non-target amplicons.

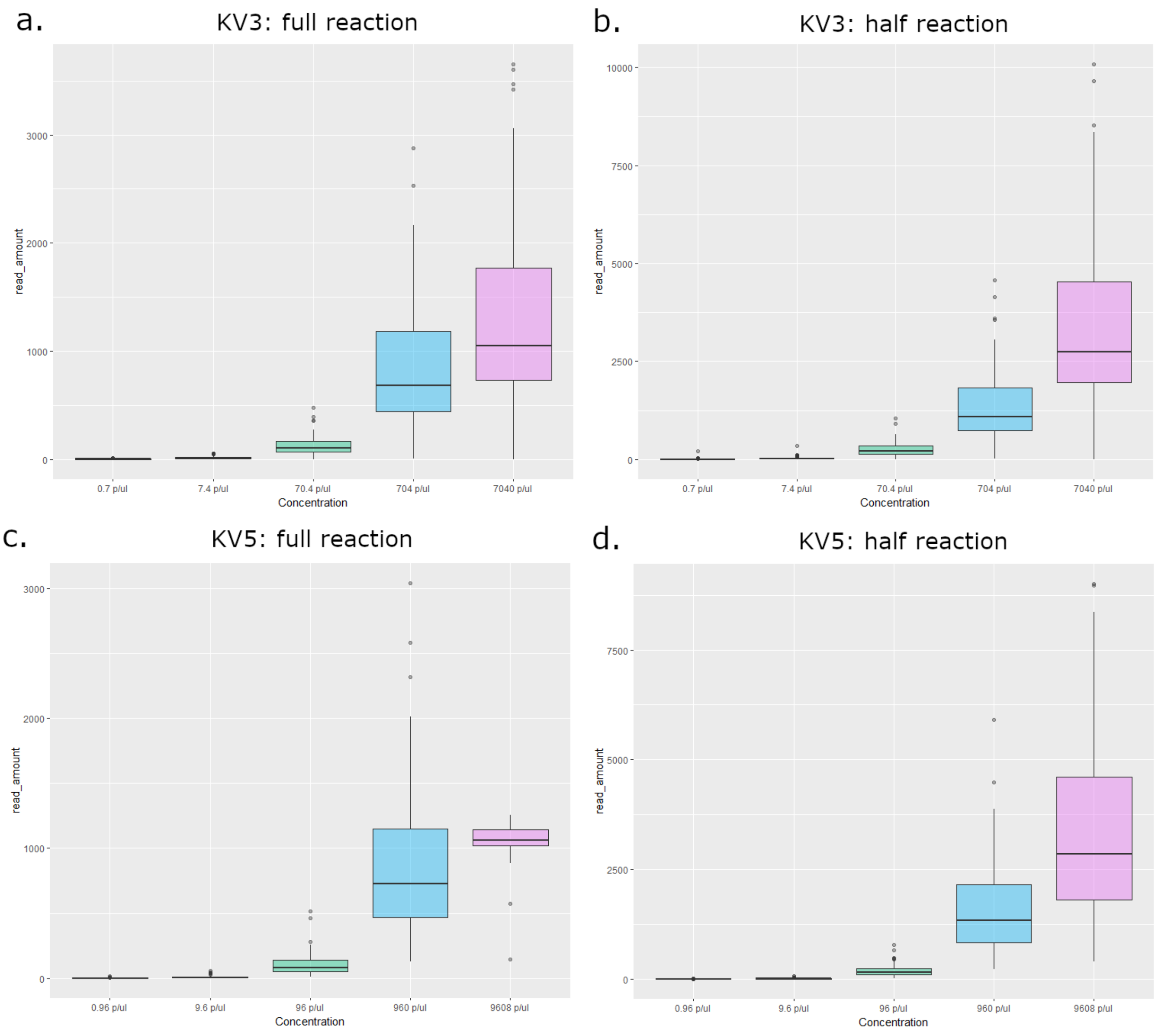

**Supplementary Figure 4. Assay sensitivity using serial dilutions.** Distributions of read counts across the 97 markers (excluding the mitochondrial marker) in two independent *P. vivax* serial dilutions, samples KV3 and KV5; in full (panels a and c) and half (panels b and d) reactions. Results reflect per marker read counts (read amount) averaged across the three replicates for each dilution. Concentration on the x-axis reflects parasite density, estimated as the number of parasites (p) per microliter (ul) of blood. Half reactions (10 ul reaction mix in library preparation PCR step 1, comprising 5.5 ul DNA) displayed slightly higher sensitivity than full reactions (20 ul reaction mix in PCR step 1, comprising 11 ul DNA). This result was unexpected and may reflect modest differences in the amount each library contributed to the final pool for the run, as the half and full reactions were pooled separately. Also of note, within each full and half reaction, the KV3 sample, with estimated 7,040 p/ul starting density, had slightly higher sensitivity than the KV5 sample, which has estimated 9,600 p/ul starting density. This trend was also unexpected and may reflect inaccuracies in the estimation of the amount of DNA in each sample as microscopy measures do not account for differences in DNA abundance between different *P. vivax* life cycle stages. Microscopy estimates also do not account for free DNA (i.e. DNA outside of the cells) or potential DNA degradation. All results reflect *P. vivax* sensitivity on 384-plex runs on a MiSeq instrument.

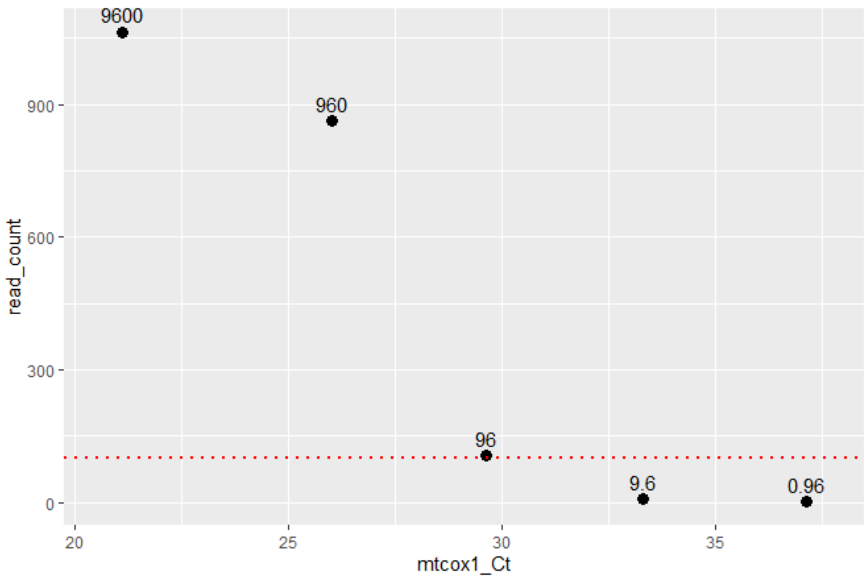

**Supplementary Figure 5. Pvmtcox1 qPCR Ct against parasite density.** Data is presented for the KV5 serial dilution, with parasite densities labeled above each point. The *pvmtcox1* Ct scores reflect an average across triplicates. The dashed red line delineates the threshold for an average of 100 reads per marker, which is reached at and above 96 parasites per microliter blood.

| Sample | Parasite density (p/ul) | Average rhAmpSeq read count[a] | Average *pvmtcox1* Ct[b] | *delta* Ct |
|---|---|---|---|---|
| KV3 | 704 | 840 | N/A | N/A |
| **KV3** | **70.4** | **125** | N/A | N/A |
| KV3 | 7.04 | 15 | N/A | N/A |
| KV3 | 0.7 | 3 | N/A | N/A |
| KV5 | 9,600 | 1064 | 21.12 | 4.90 |
| KV5 | 960 | 863 | 26.02 | 3.62 |
| **KV5** | **96** | **107** | **29.64** | **3.68** |
| KV5 | 9.6 | 9 | 33.32 | 3.82 |
| KV5 | 0.96 | 2 | 37.14 | N/A |

**Supplementary Table 1. Assay sensitivity in the P. vivax serial dilutions.** Summary of the read counts derived from the rhAmpSeq assay and cycle threshold (Ct) in the *pvmtcox1* PCR in serial dilutions of *P. vivax* samples KV3 and KV5. The KV3 and KV5 dilutions at/above which a minimum of 100 reads are yielded on average for each of the markers in the rhAmpSeq assay are highlighted in bold; below a Ct of 30-34, an average of 100 reads per marker is expected. [a] Averaged across replicates (n=3) across 97 markers in the assay (excluding the mitochondrial marker which has multiple copies per cell). [b] Averaged across triplicates.

<table>
<tr><th>10% minor allele threshold</th><th>Mhap homozygous reference</th><th>Mhap homozygous alternate</th><th>Mhap heterozygous</th><th>Mhap genotype fail</th></tr>
<tr><td>WGS homozygous reference</td><td>52.97% (17,440/32,923)</td><td>0.006% (2[a]/32,923)</td><td>0.316% (104/32,923)</td><td rowspan="3">980</td></tr>
<tr><td>WGS homozygous alternate</td><td>0.055% (17/32,923)</td><td>40.85% (13,449/32,923)</td><td>0.368% (121[b]/32, 923)</td></tr>
<tr><td>WGS heterozygous</td><td>0.844% (278/32,923)</td><td>0.149% (49/32,923)</td><td>4.444% (1,463/32,923)</td></tr>
<tr><td>WGS genotype fail</td><td colspan="3">1,318</td><td>54</td></tr>
</table>

**Supplementary Table 2. Concordance in SNP-based genotype calling between amplicon sequencing and whole genome sequencing (WGS) data at 10% threshold**. The data is derived from genotype calls derived from the VCF pipeline at 425 biallelic SNPs in 83 independent *P. vivax* samples with high quality WGS and amplicon sequencing data using the default 10% minor allele threshold. The numerator and denominator reflect the number of genotypes meeting the given criteria and the total number of successful genotyping calls across the dataset (32,923) respectively. [ab]Two and one genotype respectively with a second alternate allele.

| **1% minor allele threshold** | Mhap homozygous reference | Mhap homozygous alternate | Mhap heterozygous | Mhap genotype fail |
|---|---|---|---|---|
| WGS homozygous reference | 52.9% (17,430/32,923) | 0.006% (2[a]/32,923) | 0.346% (114/32,923) | |
| WGS homozygous alternate | 0.055% (18/32,923) | 39.89% (13,133[b]/32,923) | 1.324% (436[c]/32,923) | |
| WGS heterozygous | 0.702% (231/32,923) | 0.021 (7/32,923) | 4.714% (1,552/32,923) | 980 |
| WGS genotype fail | 1,318 | | | 54 |

**Supplementary Table 3. Concordance in SNP-based genotype calling between amplicon sequencing and whole genome sequencing (WGS) data at 1% threshold**. The data is derived from genotype calls derived from the VCF pipeline at 425 biallelic SNPs in 83 independent *P. vivax* samples with high quality WGS and amplicon sequencing data using a 1% minor allele threshold. The numerator and denominator reflect the number of genotypes meeting the given criteria and the total number of successful genotyping calls across the dataset (32,923) respectively. [abc] 2, 1 and 4 respectively genotypes with a second alternate allele.

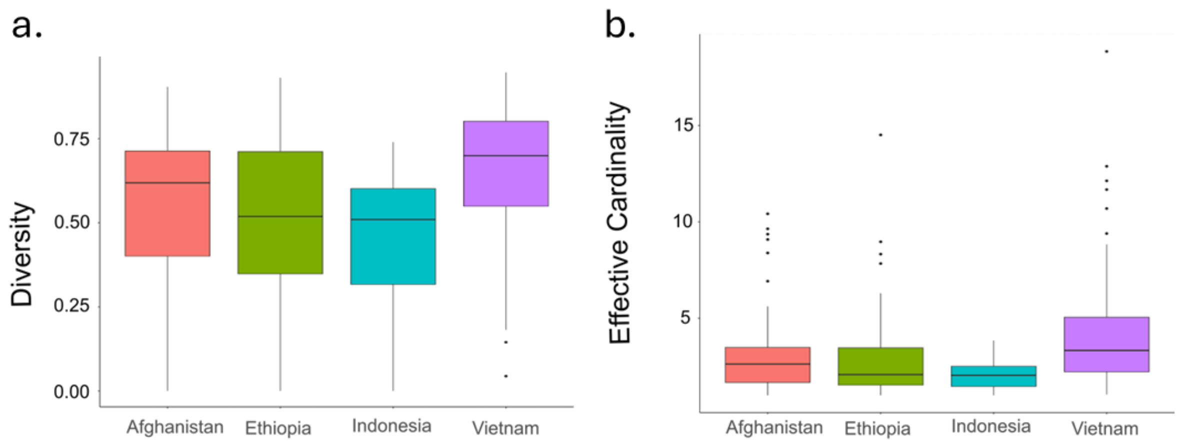

**Supplementary Figure 6. Marker diversity by country.** Panel a) presents heterozygosity measures and panel b) presents effective cardinality scores in n=498 independent samples. Each boxplot presents the median, interquartile range and min and max value for the heterozygosity (panel a) and effective cardinality (panel b).

| Sample | Run | PCR-based Species | Country | depth | coverage | Mitochondrial *Plasmodium* spp |
|---|---|---|---|---|---|---|
| QECK_1_NV22 | Run 2 | *Homo sapiens* | N/A | 2 | 0.565 | N/A |
| QECK_2_NV22 | Run 2 | *Homo sapiens* | N/A | 0 | 0.000 | N/A |
| QECK_3_NV22 | Run 2 | *Homo sapiens* | N/A | 0 | 0.000 | N/A |
| QECK_4 | Run 7 | *Homo sapiens* | N/A | 0 | 0.000 | N/A |
| RDM_47_NV22 | Run 2 | *P. falciparum* | Indonesia | 5212 | 0.996 | *P. falciparum* |
| K1 | Run 7 | *P. falciparum* | Unknown | 49,058 | 1.000 | *P. falciparum* |
| RDM_44 | Run 2 | *P. malariae* | Uganda | 56 | 1.000 | *P. malariae* |
| RDM_45 | Run 2 | *P. malariae* | Uganda | 28 | 1.000 | *P. malariae* |
| RDM_67_NV22 | Run 2 | *P. ovale* | Uganda | 150 | 0.980 | *P. ovale* |
| KK_64_NV22 | Run 2 | *P. knowlesi* | Malaysia | 2318 | 1.000 | *P. knowlesi* |
| KK_107_NV22 | Run 2 | *P. knowlesi* | Malaysia | 68 | 1.000 | *P. knowlesi* |
| KK_95_NV22 | Run 2 | *P. knowlesi* | Malaysia | 58 | 1.000 | *P. knowlesi* |
| AF001_099_D0 | Run 3 | *P. vivax* | Afghanistan | 1376 | 1.000 | *P. vivax* |
| M004 | Run 3 | *P. vivax* | Bangladesh | 158 | 1.000 | *P. vivax* |
| 05027V-1 | Run 2 | *P. vivax* | Colombia | 896 | 1.000 | *P. vivax* |

| | | | | | | |
|---|---|---|---|---|---|---|
| CQE001 | Run 3 | *P. vivax* | Ethiopia | 1058 | 1.000 | *P. vivax* |
| ID004020_D0 | Run 6 | *P. vivax* | Indonesia | 160 | 1.000 | *P. vivax* |
| MV1 | Run 2 | *P. vivax* | Malaysia | 1044 | 1.000 | *P. vivax* |
| MIDAS-129 | Run 2 | *P. vivax* | Sudan | 2158 | 1.000 | *P. vivax* |
| VN001_002 | Run 3 | *P. vivax* | Vietnam | 168 | 1.000 | *P. vivax* |

**Supplementary Table 4. Plasmodium spp. classification.** Details of the *Plasmodium* spp. Classification determined at the mitochondrial amplicon in 4 human controls, 8 independent non-vivax infections, and 8 independent *P. vivax* infections selected for representation of a range of countries. Low depth and coverage of the mitochondrial amplicon was confirmed in the 4 human controls, and concordance between PCR-based and mitochondrial classification of *Plasmodium* spp. was confirmed in all malaria infections. The number of samples in each run were as follows: run 2 (n=96), runs 3-6 (n=384) and run 7 (n=48).

| Gene | Chr | Position | Mutation | Drug | Freq, % (no./No.) Afghanistan | Freq, % (no./No.) Ethiopia | Freq, % (no./No.) Sumatra, Indonesia | Freq, % (no./No.) Vietnam |
|---|---|---|---|---|---|---|---|---|
| *pvmdr1* (PVP01_1010900) | 10 | 479908 | F1076L | CQ | 100 (159/159) | 100 (215/215) | 100 (38/38) | 92 (80/87) |
| *pvmdr1* (PVP01_1010900) | 10 | 480207 | Y976F | CQ, AQ+SP | 0 (0/159) | 41 (81/198) | 100 (38/38) | 73 (62/85) |
| *pvdhps* (PVP01_1429500) | 14 | 1270401 | A553G | Antifolate | 0 (0/159) | 0 (0/215) | 0 (0/38) | 2 (2/90) |
| *pvdhps* (PVP01_1429500) | 14 | 1270911 | A383G | Antifolate | 2 (3/157) | 10 (20/205) | 53 (20/38) | 80 (68/85) |

**Supplementary Table 5. Prevalence of orthologous drug resistance markers in each country.** Mutation prevalence was calculated with homozygous calls only. Abbreviations: Chr, chromosome; Freq, Frequency; AQ, amodiaquine; Chr, chromosome; CQ, chloroquine; MQ, mefloquine; SP, sulfadoxine-pyrimethamine.

**Supplementary Note 1. *P. vivax* rhAmpSeq Library Preparation**

Individual rhAmpSeq primers (see Appendix I) with standard desalting purification were purchased from Integrated DNA Technologies (IDT) and reconstituted to 100µM with IDTE, pH 7.5. rhAmp PCR reactions were prepared with 4x rhAmpSeq Library Mix 1 (IDT) and varied primer concentrations optimized to achieve moderately uniform depth across amplicons (range 0.2 – 5.7µM). For full reactions, a total of 11µl genomic DNA (regardless of parasite density) was included in a total PCR reaction volume of 20µl. The rhAmp PCR reaction was run using the following settings: Enzyme activation: 10 min at 95°C**;** 14 cycles of Amplification: Denaturation: 15 sec at 95°C; Annealing: 8 min at 61°C**;** Enzyme deactivation: 15 min at 99.5°C. The rhAmp PCR products were then diluted in nuclease-free water using 1 in 20 dilutions to a total volume of 100µl. Indexing PCR reactions were prepared with the following components: 2µl of nuclease-free water, 2µl of xGEN 10nt Unique Dual Index, 5µl of 4x rhAmpSeq Library Mix 2 (IDT), and 11µl of diluted PCR1 product. The Indexing PCR reaction was run using the following settings: Enzyme activation: 3 min at 95°C**;** 24 cycles of Amplification: Denaturation: 15 sec at 95°C; Annealing: 30 sec at 60°C; Extension: 30 sec at 72°C**;** Final Extension: 1 min at 72°C. Half reactions were run using half the quantity of input genomic DNA and rhAmpSeq Library Mix 1 for the rhAmp PCR, and half the rhAmp PCR product input and rhAmpSeq Library Mix 2 for the Indexing PCR reactions. The thermocycling conditions for the half reactions were the same as for the full reactions.

After amplification, we pooled up to 384 samples with 5µl of each individual library combined in a 1.5mL LoBind Microcentrifuge tube. The pooled libraries were purified to remove primer dimers using 0.7X SPRI beads (Beckman Coulter). After incubating for 10 minutes at room temperature, a magnetic rack was used to separate beads and remove the supernatant. Beads were washed twice with freshly prepared 80% ethanol and left to stand for 3 minutes to enable any leftover ethanol to evaporate. The library (of preferred interest size) was then eluted in 22µl of IDTE, pH 8 (IDT). The success of library purification was evaluated by running pre- and post- bead cleanup libraries with capillary electrophoresis using an Agilent 4150 TapeStation system using D1000 reagents and ScreenTape. The pooled libraries were quantified using Collibri™ Library Quantification Kit and diluted accordingly to 4nM concentration. The diluted library was the sequenced on an Illumina MiSeq platform at 9pM final loading concentration with 10% PhiX. Sequencing was conducted using the Illumina v2 kit following the protocol for paired end 150 bp reads.

### Appendix I - *Plasmodium vivax* rhAmpSeq Primer Sequences

| AssayID_IDT | Chr | Start | End | Target | *Marker type | PrimerSequence_FWD | PrimerSequence_REV |
|---|---|---|---|---|---|---|---|
| RH.9CAFEBAA3F384A0Z0Z | 1 | 395355 | 395521 | 7907 | Mhap | /rhSeq-f/ACN CCC CAA ATG TGA ATA ArCT TCC /GT4/ | /rhSeq-r/ACG TGG CTA CTA CCC CArG TGG T/GT3/ |
| RH.2960ABE5E62C470Z0Z | 1 | 483735 | 483883 | 9674 | Mhap | /rhSeq-f/TCC AAA CTN AGC TCC TTG ATrG TTG T/GT3/ | /rhSeq-r/CAA CTT TGG CAT CCT CTA TAA CArC GGA T/GT1/ |
| RH.B2905CD19921440Z0Z | 1 | 612225 | 612348 | 12243 | Mhap | /rhSeq-f/ACC TGG AAA CTC CCT TGT TrGC AAT /GT2/ | /rhSeq-r/ATA CGA ATT CGC ATC AGA CGrG AGA G/GT4/ |
| RH.79D714EC610C448Z0Z | 1 | 772093 | 772211 | 15441 | Mhap | /rhSeq-f/TCC GAA CCA TCG CTG TTA rCCA CT/GT2/ | /rhSeq-r/CTG CCC CTT TCT CCA GAG rCAG TA/GT2/ |
| RH.964D47B0B088468Z0Z | 2 | 213958 | 214133 | 24710 | Mhap | /rhSeq-f/ACC TGG AAT GCT CCA AAA ATT rCCT TG/GT1/ | /rhSeq-r/GGT AGT GTA CAG GGA AAT CAC rCCC GA/GT1/ |
| RH.AC2609A2062C4D4Z0Z | 2 | 327552 | 327739 | 26982 | Mhap | /rhSeq-f/TTC GCA ACA AGA GGA GCA AArC ATA G/GT2/ | /rhSeq-r/CCG TCA AAT GGT AAA GCG TrGA AGN /GT4/ |
| RH.C0DE1AD20EB44E6Z0Z | 2 | 660463 | 660614 | 33640 | Mhap | /rhSeq-f/GCA GCG CAT GGA AAG TAT TGrC TAG A/GT3/ | /rhSeq-r/TGA TCC ACT GCC TTT TGG TAG rCAT TC/GT3/ |
| RH.ADD86C5C7CEE4FBZ0Z | 3 | 178937 | 179094 | 43133 | Mhap | /rhSeq-f/TGG CAT AGC TGC GAA GTT rATT CA/GT3/ | /rhSeq-r/NTC CAC GTG GCT GTA TrAG GGG /GT2/ |
| RH.AC6AF27F13E1456Z0Z | 3 | 252061 | 252170 | 44595 | Mhap | /rhSeq-f/TTC GAT TTG GAA TCC CCT TrCT GCN /GT4/ | /rhSeq-r/CAA GAA AAC CCC ACC TTT GrCA CAA /GT1/ |
| RH.7702F5B5FFEA486Z0Z | 3 | 536620 | 536711 | 50287 | Mhap | /rhSeq-f/TCC CTG CTG AAG GAC TCrC GAG C/GT1/ | /rhSeq-r/ACT CAC CGN CAA CGT TrGG GCG /GT3/ |
| RH.10A2E23284CC46CZ0Z | 3 | 763605 | 763660 | 54827 | Mhap | /rhSeq-f/CTC CTG GCA TGG ACC CrCA CCT /GT3/ | /rhSeq-r/TAN AGC AGG CGG TAG AGrC TTC C/GT1/ |
| RH.00A0825CCA024A4Z0Z | 4 | 361787 | 361899 | 64721 | Mhap | /rhSeq-f/GAC CAA AGA GGA GAA AAC GArA AAA C/GT4/ | /rhSeq-r/TCC TCT TTC ACC TGC TCG rCAT GT/GT3/ |
| RH.9A9B6B1CE8ED464Z0Z | 4 | 500160 | 500273 | 67489 | Mhap | /rhSeq-f/CAT GAG GTA GTA GCT CTT CGA rCGA GT/GT1/ | /rhSeq-r/GCT TCC CCA TGG AGG GrCC TCC /GT3/ |
| RH.FC72753AD141415Z0Z | 4 | 853861 | 853944 | 74562 | Mhap | /rhSeq-f/CTG CNC AGT TTG ATC AGT CrCA CCC /GT1/ | /rhSeq-r/CTT CAT TAT TTC GAA TGG CTT TCT rGGA AG/GT1/ |
| RH.FDE3FDC2B0774C8Z0Z | 5 | 170476 | 170547 | 81132 | Mhap | /rhSeq-f/TGA ATC CTC CGA AAA CGA TTC rCTC AG/GT3/ | /rhSeq-r/GCA GTC TGA AGA TTC TGA TGrA AGA A/GT3/ |
| RH.34E5DDB83D7D4F7Z0Z | 5 | 490198 | 490244 | 87526 | Mhap | /rhSeq-f/CCC TCA TCA ATC ACT TCT TCC TArC AGA A/GT2/ | /rhSeq-r/CTT TTG CGC AAA TAA ATC CAA GTrG AAA C/GT4/ |
| RH.34F85DB3B3264F2Z0Z | 5 | 722013 | 722089 | 92164 | Mhap | /rhSeq-f/GAC GAG CAA ATT TAA GAA GCT CTC rGTA GC/GT1/ | /rhSeq-r/CTG CCA CAT CCT AAA TCA CAT ACT TrCA TAA /GT4/ |
| RH.6A1D6D2210DF49BZ0Z | 5 | 776828 | 776973 | 93261 | Mhap | /rhSeq-f/ATG AGA TTC ACA CTG TAG TCG GrGG CAG /GT2/ | /rhSeq-r/CCT CGT ATC GTT CCT TNA GTC rCTC TT/GT4/ |
| RH.83D956F8758C4BBZ0Z | 5 | 1125680 | 1125748 | 100236 | Mhap | /rhSeq-f/CCA CGC AGA GTG CTT TTrC CAT C/GT1/ | /rhSeq-r/CTT GTC TCA CCG CTG CrCC TCA /GT3/ |

| | | | | | | | |
|---|---|---|---|---|---|---|---|
| RH.42DF2C222BA74C2Z0Z | 5 | 1352101 | 1352165 | 104765 | Mhap | /rhSeq-f/GGT TCG ACA TTA TGA GTA GAC ACrG TTT G/GT4/ | /rhSeq-r/CGT TGA CCT TTT GGG AAA CAT ArCA CAT /GT1/ |
| RH.46D4266C768744DZ0Z | 6 | 153460 | 153574 | 111288 | Mhap | /rhSeq-f/CAA TTT TGC GAG GGC TAT TCrC GCA C/GT1/ | /rhSeq-r/TCA CAT GAA GTG TGC AGT TrGC TGG /GT2/ |
| RH.5312BCFBF8D8457Z0Z | 6 | 272690 | 272815 | 113672 | Mhap | /rhSeq-f/TTA GAA GTC AAT GCG ACG CrCA GAT /GT1/ | /rhSeq-r/TCT GAA TGA CCT TCC GGA rGCT GG/GT2/ |
| RH.B21F2DF1DA6A419Z0Z | 6 | 531256 | 531393 | 118844 | Mhap | /rhSeq-f/AGT CCT GCT CTC AGG GrGT CCT /GT4/ | /rhSeq-r/AGG ATG CTC ACC AGG CrGG ACA /GT2/ |
| RH.4133F987D8A44E5Z0Z | 6 | 983157 | 983333 | 127882 | Mhap | /rhSeq-f/GGA TAT GGA AGG CAN CGG ATrA TTC C/GT4/ | /rhSeq-r/TAA TCC CTT CCC CAT TCT CGrA ATC C/GT4/ |
| RH.50720784FEED46AZ0Z | 7 | 78143 | 78172 | 130632 | Mhap | /rhSeq-f/GTT CTT TTA AAT AAT GCA CCT TTT TCG rCCA TC/GT1/ | /rhSeq-r/GAA AAC CAA AAT AGA TGA AAG TTT ACA AArC AGT G/GT2/ |
| RH.3E77B333C8DB4D2Z0Z | 7 | 496520 | 496613 | 139001 | Mhap | /rhSeq-f/AGA ATG TGT CGG ATT TTC GAT TAG rGAC TC/GT1/ | /rhSeq-r/ACA ACC GCA TGT ACA ATC TTT TrGA AGG /GT4/ |
| RH.DEF0990049A641CZ0Z | 7 | 697428 | 697579 | 143020 | Mhap | /rhSeq-f/CTC ACT CAT GGA TGG GTA CAT AGrA AAA C/GT4/ | /rhSeq-r/GGG AAC CAC ATT TAC AGA TTA TCA ArAT GAG /GT2/ |
| RH.0AB7872636F3460Z0Z | 7 | 832233 | 832371 | 145716 | Mhap | /rhSeq-f/TAC ACC CNT TCG TTT AGC CrAT TTG /GT4/ | /rhSeq-r/TGA TGT AAT CCC CTG CAC AGrC TCT G/GT4/ |
| RH.C03EF5B621FF4C0Z0Z | 7 | 1123202 | 1123330 | 151535 | Mhap | /rhSeq-f/CGA GAT GTA AAC GAA GGT GArA AGG G/GT2/ | /rhSeq-r/AGA CTC ACC AGA TTG ACC ArGA CTC /GT4/ |
| RH.CBFE7DBF5A6E4D8Z0Z | 7 | 1218911 | 1219036 | 153449 | Mhap | /rhSeq-f/GAG ATT TTG CTG AAG TAC TAT AAG GrCA CGA /GT2/ | /rhSeq-r/GAT AAT TTC CTT CAG CTC TGT CAA rGAC GT/GT2/ |
| RH.0A6CDC396639498Z0Z | 7 | 1427121 | 1427291 | 157614 | Mhap | /rhSeq-f/GGT AGT CGC AAA GAA CAC TrCA NGT /GT1/ | /rhSeq-r/CAG GAA ATT TGG AAA CGC CArG TAT G/GT3/ |
| RH.82A3E1ACDE394D8Z0Z | 8 | 168547 | 168665 | 165484 | Mhap | /rhSeq-f/TGT TCC TCA CTT CTG AGA GTrA GAA G/GT3/ | /rhSeq-r/GAC CAA GTG ACG AAG CArC TAC A/GT4/ |
| RH.CC11B8DE53FA43DZ0Z | 8 | 822406 | 822480 | 178560 | Mhap | /rhSeq-f/AAA CCG AAC GTT TTA AAT GGG rCAC GT/GT2/ | /rhSeq-r/GAC AGA ACC CAC TCG TAT ATC rCCA TT/GT2/ |
| RH.4166D3F17C374C4Z0Z | 8 | 1018073 | 1018248 | 182475 | Mhap | /rhSeq-f/CCT CGA GAA GGC CAT AGT GrAG CAT /GT3/ | /rhSeq-r/GAG AGG GTC ACC GGG TrCT AAG /GT3/ |
| RH.6818F263362044BZ0Z | 8 | 1157514 | 1157634 | 185264 | Mhap | /rhSeq-f/AGA GGC TCC TAA AAG TGC TTrG TTA A/GT1/ | /rhSeq-r/GTG GGT ACT CCT CAA GTG TTT rAAT AT/GT1/ |
| RH.CD3AC424DC4D417Z0Z | 8 | 1403792 | 1403883 | 190188 | Mhap | /rhSeq-f/CAC GAA TAC ATG CAT GTG TGT rGCG CA/GT1/ | /rhSeq-r/TCG TCG TCG TTA TGT ATG CTG rCAG TC/GT2/ |
| RH.D5E3F1910FBA4C7Z0Z | 8 | 1468975 | 1469148 | 191493 | Mhap | /rhSeq-f/GAG ATC ACC AGA CCA CAG rGAG CA/GT2/ | /rhSeq-r/CTG CAT TCA TGT CNT TCG AAA ArAT TGT /GT1/ |
| RH.1B258A1478D74CDZ0Z | 9 | 264626 | 264698 | 202627 | Mhap | /rhSeq-f/GTT TGA GGA AAA TCT CGA AAG AAG AArC TAG C/GT3/ | /rhSeq-r/TTG CTT ATC TCA GCA CTG CTT TrGG TCA /GT3/ |
| RH.36E8980535C54B7Z0Z | 9 | 448574 | 448630 | 206308 | Mhap | /rhSeq-f/CCA GCT GTT TAT TTT CAA TCA AGT rGGT GA/GT1/ | /rhSeq-r/CAC GAG AAA AGA AAA CGA AAT TGA rAAG CT/GT4/ |
| RH.51F901C8B81F436Z0Z | 9 | 756312 | 756407 | 212462 | Mhap | /rhSeq-f/CAT GCC CAC GCA GGT ArCA CTC /GT4/ | /rhSeq-r/CAC TCT CTN CAT GGG ATA ACT rAAA AC/GT2/ |

| | | | | | | | |
|---|---|---|---|---|---|---|---|
| RH.C5069931FF214E6Z0Z | 9 | 842507 | 842645 | 214187 | Mhap | /rhSeq-f/TAA CCT CTT CAG CAT GAG AGT rCAT CG/GT4/ | /rhSeq-r/CCA ATC GAA AGG TTG GCC ArCT TTA /GT4/ |
| RH.290757F70A964E4Z0Z | 9 | 1165616 | 1165689 | 220647 | Mhap | /rhSeq-f/TTT GCC TCC CTA CTT GAA rGTA CG/GT2/ | /rhSeq-r/CGA CAT TAA CCT GAA CAC CTrG GTC A/GT4/ |
| RH.BD4EC483E885416Z0Z | 9 | 1459600 | 1459729 | 226528 | Mhap | /rhSeq-f/GTC CTG TTT TTG GAA AGG GTrA TNG T/GT3/ | /rhSeq-r/CAA AGC TAG CTG CGT GGrG TTC T/GT2/ |
| RH.52CD33BD1DC7471Z0Z | 9 | 1688611 | 1688696 | 231108 | Mhap | /rhSeq-f/AGC AAG GAC AAG ATG AGG ATrG AAC A/GT4/ | /rhSeq-r/GCA TTT AAG GAC ATG CAA CTG rGAA TC/GT4/ |
| RH.E03EC290DCDC420Z0Z | 9 | 1864594 | 1864642 | 234628 | Mhap | /rhSeq-f/CAT CAT CAC ATA TGC TAT CAT TGT rCTG CC/GT4/ | /rhSeq-r/ACA AGT ACA AAA CGA TGA GCA AAA rGTG GA/GT2/ |
| RH.E7EBCAE148AF44CZ0Z | 9 | 2150871 | 2150956 | 240353 | Mhap | /rhSeq-f/CTT CTG AAT TTT TCA TAA ATT CAT CCC rATT GC/GT1/ | /rhSeq-r/GAG ATG GTT TAC CTT CAC TTC TrCA ACC /GT4/ |
| RH.F6F8C3D0644D440Z0Z | 10 | 203316 | 203481 | 246142 | Mhap | /rhSeq-f/GCT CAC TTG GTT TCT TTT TAC CArG AAT G/GT2/ | /rhSeq-r/AGG AAA GCA ACA GGG CAT rCCN AA/GT4/ |
| RH.5B17C2D0F468424Z0Z | 10 | 254364 | 254412 | 247163 | Mhap | /rhSeq-f/GTT TTA CCA GAA TAT GTG GAG CAT rGAA AG/GT4/ | /rhSeq-r/GCG TAT AGG ATC TGA ATA GTC ATC GrAT TAG /GT4/ |
| RH.0D4A6DF10E97412Z0Z | 10 | 479907 | 479908 | MDR1_F1076L | Drug | /rhSeq-f/TTT AGG GAC ATC AAC TTC CCG rGCG TA/GT2/ | /rhSeq-r/AGA CGC TAA TAA ATT CGA TGC TrCT GGG /GT2/ |
| RH.45AE3D12F5854F9Z0Z | 10 | 480206 | 480207 | MDR1_Y976F | Drug | /rhSeq-f/TTC TTC TCT ACA TCC TTG TTG GrCT GCT /GT4/ | /rhSeq-r/CTC ACT TTA TAG TGC TCT TCC TTrG TGA G/GT1/ |
| RH.DEAB619F5DDE469Z0Z | 10 | 725009 | 725169 | 256576 | Mhap | /rhSeq-f/GCT GTT GAT ATC AAA TGT GCT rCGT CC/GT4/ | /rhSeq-r/AAG AAG AGC AAG AAG GAG TTrC ACC C/GT4/ |
| RH.C93D7255B094471Z0Z | 10 | 848868 | 848916 | 259053 | Mhap | /rhSeq-f/GTA AAA CTG TTT GAT ATC CCC GTT rGGT TA/GT4/ | /rhSeq-r/CTT GGA AAG ACA ACA AGA AAC ArCG GAG /GT2/ |
| RH.D97789F3C3B0468Z0Z | 10 | 1097783 | 1097940 | 264031 | Mhap | /rhSeq-f/GAT GAA TTC ATC CGT TTG GCrG ATG G/GT4/ | /rhSeq-r/TGN AAA AGC TAA ACA TCC TAA ACrG AGC T/GT3/ |
| RH.32854966B3C54A7Z0Z | 10 | 1271498 | 1271644 | 267505 | Mhap | /rhSeq-f/GCT AAT GTC TCT ACT AAC GTC TCT rACT AA/GT3/ | /rhSeq-r/CAC GCA GGA GGC AAA NTA TrCA TTT /GT2/ |
| RH.25183DF8EA034DAZ0Z | 10 | 1408100 | 1408170 | 270238 | Mhap | /rhSeq-f/GCA AAA TGA TGA GTA TTC CAT GAT TTT rCTG TG/GT1/ | /rhSeq-r/GTA AAA AGG ATG CTC ATT TTG CTrG CAG G/GT1/ |
| RH.6C64A045C9CA4D5Z0Z | 11 | 75770 | 75798 | 274564 | Mhap | /rhSeq-f/CCA ATT TAT GGT AGA GGA TTA GTA TCrA CTT G/GT3/ | /rhSeq-r/CTT TAC TAA TTT CAG TTA TGT ATA ATG CCrC ATT G/GT1/ |
| RH.3909CDB67E8543CZ0Z | 11 | 177898 | 177971 | 276606 | Mhap | /rhSeq-f/GGT TTT CAC TCC CTC CAC TrCA TTT /GT1/ | /rhSeq-r/GGC ACT CTT TTG AGT AGC AGrC TTG A/GT3/ |
| RH.E13F9A10B62A4BEZ0Z | 11 | 607788 | 607927 | 285205 | Mhap | /rhSeq-f/TAA CCA CCA CTG TGT TAT CCA TAT rCTG TT/GT1/ | /rhSeq-r/TTA ATA AAG ACA CNA ATG TAG ATT TGA ACrA ACA T/GT3/ |
| RH.83B95CBC9B044D2Z0Z | 11 | 916515 | 916676 | 291380 | Mhap | /rhSeq-f/GCA CTN TCT GAT AGC ATG TrGG TCT /GT4/ | /rhSeq-r/GAG ACC CAT CCA CAT CTG rCGA AT/GT4/ |
| RH.7D6C9D6F4F5044AZ0Z | 11 | 1153666 | 1153850 | 296123 | Mhap | /rhSeq-f/ATG TGA CGT CTC TCC ACC rCCC CT/GT2/ | /rhSeq-r/CTA CAT CCA NCA TAC TCT GCrA GGT A/GT1/ |
| RH.230B0FFB8743402Z0Z | 11 | 1423369 | 1423519 | 301517 | Mhap | /rhSeq-f/CAA AGT TGT AAA AGN GAT CTG CTC ArAA TTT /GT3/ | /rhSeq-r/CAA GCC TGA CTG TTC AGA AAA ArAT TTC /GT1/ |

| | | | | | | | |
|---|---|---|---|---|---|---|---|
| RH.9D2EBAFECC02434Z0Z | 11 | 1572690 | 1572840 | 304503 | Mhap | /rhSeq-f/GCC GCT CTA CAA GGG ArGA AGT /GT2/ | /rhSeq-r/CCT TGC GCC TGA AGT TAT rCGT AC/GT1/ |
| RH.2449A198108D4EDZ0Z | 11 | 1742120 | 1742207 | 307891 | Mhap | /rhSeq-f/GTT GTT AAC TCG TAA GCT GTT GArG GAA G/GT3/ | /rhSeq-r/AGG CGA ATA ACC CAC GTA AGrG ACA A/GT3/ |
| RH.DC722708FE7E46FZ0Z | 11 | 2001515 | 2001699 | 313080 | Mhap | /rhSeq-f/GCN CTT CAT GTT TAC AGT GTrA AGC A/GT3/ | /rhSeq-r/TCA TTC GCC TCG ATG GAA rGAC AC/GT2/ |
| RH.B289254C8C0F4D9Z0Z | 12 | 324220 | 324242 | 322153 | Mhap | /rhSeq-f/CTG GTG AAG GTG AAG GAA AAT GAA ArCT ATT /GT3/ | /rhSeq-r/CCA TCT ATG TTT TCC GTT TTC TGC rGTC TT/GT3/ |
| RH.B058277019D04D7Z0Z | 12 | 532970 | 533101 | 326331 | Mhap | /rhSeq-f/GAA ACG AAA TAT GCC GAC ATC TrAT TCT /GT1/ | /rhSeq-r/TTG CAA AAA GCC GAA GGT TTT rCAC CA/GT2/ |
| RH.64D26ADFAE5B4C9Z0Z | 12 | 671183 | 671280 | 329094 | Mhap | /rhSeq-f/AGT GGA ATT TGT AAA AAT ATT AAG TAT GArA GTA C/GT1/ | /rhSeq-r/CTA TCA AAC ATG TCA ACG ACT GAA GrGA GAT /GT4/ |
| RH.AB04234C54EB428Z0Z | 12 | 995697 | 995758 | 335584 | Mhap | /rhSeq-f/CAT ATG TGT TTG AAC GAT TCT TAC GTrA TGT T/GT1/ | /rhSeq-r/TGG AAA AGC GAT TCA TAA TTT TTA GAG rCAA CG/GT1/ |
| RH.F5FE56DD7B144F1Z0Z | 12 | 1291327 | 1291407 | 341498 | Mhap | /rhSeq-f/CTT TAT GTT GGA GAC TGA TTT GTT rCGC CT/GT1/ | /rhSeq-r/GTG CTA ATC GAG AAG ATC CTA ACrG AAC G/GT3/ |
| RH.D427FB9C888748FZ0Z | 12 | 1457399 | 1457514 | 344819 | Mhap | /rhSeq-f/GAG AAT AAA ATA CCC CTT CAA ATG GArG GAA G/GT1/ | /rhSeq-r/CAT ATG TCA ATG TGA TTA CTT CTT GGT rGAA GC/GT4/ |
| RH.E926E30943B14DCZ0Z | 12 | 1880553 | 1880581 | 353283 | Mhap | /rhSeq-f/GCT TGG TTT TCT GCA CCA GrGT CAT /GT3/ | /rhSeq-r/TTG GAC GCC TTC CTG AAG rGAC TC/GT3/ |
| RH.7B5B62F5A6C54CBZ0Z | 12 | 1945902 | 1946055 | 354590 | Mhap | /rhSeq-f/TGT ATN CTC CAT TTG GGA GTrC CAC C/GT1/ | /rhSeq-r/CCA GAT GAG CAA CCC AArC GGA A/GT2/ |
| RH.A4206DB085EE47DZ0Z | 12 | 2108485 | 2108611 | 357841 | Mhap | /rhSeq-f/TGA TGG TCT TTA CAC ACT CGT rATA GG/GT4/ | /rhSeq-r/AAC CCG ATG ATG CCA TAG AArG GTG T/GT1/ |
| RH.524A823035CE473Z0Z | 12 | 2465991 | 2466085 | 364991 | Mhap | /rhSeq-f/CCA TAT TTA TAT CTT CAT CAT CGC TTT TrCT TCC /GT3/ | /rhSeq-r/ATT ATA AAT TCA GAC TCA TCT TAT TCA TCrC NAT G/GT4/ |
| RH.8971DA4524DB42FZ0Z | 12 | 2724027 | 2724063 | 370151 | Mhap | /rhSeq-f/TCA CTC CTT TGC TCA GTC rCTG TT/GT4/ | /rhSeq-r/GAC GAA ACG GAT AAA TTG CTA CArC TAC T/GT4/ |
| RH.D7309B8EBCBD4ECZ0Z | 13 | 167460 | 167596 | 382674 | Mhap | /rhSeq-f/TGG ACG GCG ACA TCT GTrG AAC T/GT1/ | /rhSeq-r/CGG AGC TGT TTA GCA GGT rCAT TC/GT2/ |
| RH.F252700317D4403Z0Z | 13 | 293615 | 293708 | 385196 | Mhap | /rhSeq-f/AAC TGT CTC AGG TAA TTG CCrC CCT C/GT3/ | /rhSeq-r/AGT TGG AAA GGA GAC AGA AAA ATrA TGG C/GT4/ |
| RH.9E1CA1645B3C421Z0Z | 13 | 485702 | 485833 | 389038 | Mhap | /rhSeq-f/GTG GGA TGG TCT CTA CTT ATrG TGA C/GT3/ | /rhSeq-r/CAA TAA CAG CTC CTT CAA CTT rCGA GA/GT4/ |
| RH.CA5B246F7CB148AZ0Z | 13 | 754813 | 754885 | 394419 | Mhap | /rhSeq-f/GAA GTG TTA AAA TTA ACT GGA GCA ArAT ATG /GT4/ | /rhSeq-r/GCA CAT CAT TGT AAT CCT GGA TrGA AGG /GT4/ |
| RH.49DD6A84FF2848AZ0Z | 13 | 1197713 | 1197831 | 403278 | Mhap | /rhSeq-f/GCC GAC TAT CGC ACT TTT rGTT CT/GT2/ | /rhSeq-r/TTG CAG AAG GAT GCT CTG ArAT GAG /GT4/ |
| RH.28A3310975E14FDZ0Z | 13 | 1865343 | 1865483 | 416631 | Mhap | /rhSeq-f/TTT CCG TGG CTN AGT GGrC GAC T/GT1/ | /rhSeq-r/AGG TGT CAG CGC TAG CrGG CAG /GT1/ |
| RH.668CC46600DF42AZ0Z | 13 | 1985698 | 1985814 | 419038 | Mhap | /rhSeq-f/AAG AAA CTG CTA TAC TGT TTG CrCT ACG /GT4/ | /rhSeq-r/ATC CAT TGA ATA ACC CGC TTT rGCA CT/GT3/ |

| | | | | | | | |
|---|---|---|---|---|---|---|---|
| RH.AA0D417E764C4FDZ0Z | 14 | 56852 | 57021 | 422331 | Mhap | /rhSeq-f/CAA TAA ATC GAC CAA GNT TCT TTC CArG AAT A/GT4/ | /rhSeq-r/TGC ATT TAT CTA TTA TGG TAG CAT AAT GGrA TAA T/GT4/ |
| RH.EA14E24E533148FZ0Z | 14 | 462743 | 462837 | 430447 | Mhap | /rhSeq-f/CAG GCT CAT TGG AAT GGT TGrC TAC T/GT2/ | /rhSeq-r/AAT ACA GAA GTG TAC CAA GCC rGTA GC/GT1/ |
| RH.D19C9EEADCE7431Z0Z | 14 | 675630 | 675699 | 434704 | Mhap | /rhSeq-f/AAC GAC ATC CTC AAT TGG AAA rCAG GG/GT2/ | /rhSeq-r/GTC CCA AAC TTT CAA GCT GTrA AAA G/GT1/ |
| RH.FF5E58E56C4B4E5Z0Z | 14 | 1206462 | 1206642 | 445323 | Mhap | /rhSeq-f/CTT ATT GTG CAG GGA AAA CCA rCAA AT/GT4/ | /rhSeq-r/TCA GGG AGC TAA ACG ATT ACA rGCA AC/GT1/ |
| RH.2CD34BE4DF1F4B6Z0Z | 14 | 1270400 | 1270401 | DHPS_A553G | Drug | /rhSeq-f/CTG CAA CAG CTT AAT AGA CTG rGTC GT/GT2/ | /rhSeq-r/AGC GTC GTT TTA ATG CAC AArG AGG G/GT2/ |
| RH.C975D06C896447DZ0Z | 14 | 1270910 | 1270911 | DHPS_A383G | Drug | /rhSeq-f/AAC CTC ACA CTC CAA CTT ATG rCCA CT/GT3/ | /rhSeq-r/CTT TTC AGA TGG CGG TTT ATT TrGT CGA /GT1/ |
| RH.436418B818FD4C7Z0Z | 14 | 1546237 | 1546249 | 452117 | Mhap | /rhSeq-f/ATT GGG AAA CAG GAG AAA TGT TTA TrGG GTA /GT1/ | /rhSeq-r/TAA GTC GTA ACC ATC AGG TAG TTT TrAT GAA /GT1/ |
| RH.21577148368744AZ0Z | 14 | 1763089 | 1763164 | 456454 | Mhap | /rhSeq-f/CGC ATG CAA AAG GAA AAT TAA ATG TTrC ACT C/GT3/ | /rhSeq-r/ACA AGA GTT ACA CTA TTC GCT TTT rGCG CT/GT3/ |
| RH.04C2CDE0F619407Z0Z | 14 | 1887637 | 1887677 | 458944 | Mhap | /rhSeq-f/GTT AGA GAG TGG CAT GGA TGT rGAA TT/GT1/ | /rhSeq-r/AAT ACC CGA GCA TCC TAA ACA rGAT CA/GT4/ |
| RH.771A53FD18D3457Z0Z | 14 | 2022558 | 2022663 | 461645 | Mhap | /rhSeq-f/TCC CTT TTC TAT GAG GCT AAC TArG CTC T/GT3/ | /rhSeq-r/GGC AAC GAA CTC ATC CAA TTA GrGA AAC /GT2/ |
| RH.33061656A3E54DDZ0Z | 14 | 2189680 | 2189767 | 464987 | Mhap | /rhSeq-f/CCT GCA GTT TGC CTT TTT GrCA CAT /GT3/ | /rhSeq-r/AGA ACC TCC ACG CAG TAC rCTG TT/GT1/ |
| RH.0473747F9CDA4F7Z0Z | 14 | 2261604 | 2261794 | 466426 | Mhap | /rhSeq-f/ACT ATC TAA CGA GTA GCA GCrA GCG G/GT1/ | /rhSeq-r/CGT TGT TNA GAG GTC CTC TrCT GGA /GT2/ |
| RH.3AE56662E23E463Z0Z | 14 | 2699925 | 2700045 | 475191 | Mhap | /rhSeq-f/AAA ACG GTG CTC TTG TCG rGTG GT/GT4/ | /rhSeq-r/GAA AAA GTG GGC CCG GTrG GAA A/GT2/ |
| RH.BDE839E951D14CAZ0Z | 14 | 2861965 | 2862134 | 478433 | Mhap | /rhSeq-f/AAC TTA TTG AGG ATG TTA TGG AAG rAGN GA/GT4/ | /rhSeq-r/CTT ACT GCA CCC AGA CAA TAA GrGA CTT /GT2/ |
| RH.B3F556D4B68B487Z0Z | 14 | 3009930 | 3010081 | 481392 | Mhap | /rhSeq-f/GCG ATA TCA CTT TTT AAG TCA TCG rAAT GA/GT4/ | /rhSeq-r/TAA GCC ACT AGN GTA TGA TGA CGrA ATT T/GT3/ |
| RH.7E13E7FC3A8D47CZ0Z | MIT | 2930 | 3141 | MIT_Species2 | Species | /rhSeq-f/CAT CGC AGC CTT GCA ATA AAT TAA TrAT TAT /GT1/ | /rhSeq-r/CAG TCG AGT TCC TTT AAT GTA GTT TCrC TCA C/GT4/ |

*Mhap (microhaplotype), Drug (putative drug resistance marker), Species (mitochondrial *Plasmodium* spp. marker).

**Supplementary Note 2. Bioinformatic Pipeline for rhAmpSeq data analysis**

The snakemake-based pipeline provides two separate data analysis options that both require demultiplexed fastq files in a separate folder as input. As part of the pipeline, parameters are given in the config.yaml file, which can be modified as required.

a) The first analysis option performs SNP based variant calling using the provided reference genome PvP01_v2. The steps are as following:

(1) Generate manifest: This traverses the directory, identify the input fastq files and create a metadata file. This ensures that the forward and reverse reads belonging to the same sample are treated as such. The manifest is subsequently used by the pipeline to place the reads of each sample into its own directory adhering to the directory structure the pipeline requires.
(2) Mapping: The fastq files are mapped against the provided reference genome using bwa-mem2 and can be optionally trimmed prior. The alignment map (BAM) files are subsequently filtered to reads mapped in proper pairs.
(3) Base calibration: This step involves generating a recalibration table to detect systematic errors made by the sequencing machine when estimating the accuracy of each base call by comparing them to known sites of variation using GATK's BaseRecalibrator. The base quality scores in the input BAM file are then adjusted with GATK's ApplyBQSR according to the patterns identified in the recalibration table. The known variant database can be updated with knownvariants_dir in config file.
(4) Individual sample variant calling: Variant calling is performed for each sample against the reference genome using GATK's HaplotypeCaller with the following parameters "--max-reads-per-alignment-start 0 --do-not-run-physical-phasing --pileup-detection --dont-use-soft-clipped-bases". The parameters can be tweaked at the config file under haplotypecaller_flags.
(5) Joint variant calling: The GVCF outputs from previous step are used perform joint variant calling with GATK's GenotypeGVCFs. This step generates the final VCF outputs containing all the variants found in all the samples.

b) The second analysis option creates microhaplotypes from the same input files using 1) fasta files containing the forward and reverse primer sequences for all target markers, (2) a bed file containing the chromosome, start position, end position and name of each target marker, as well as (3) a fasta file of the target regions from the PvP01.v2 reference. The individual processes are listed here:

(1) Trim: Trimming of adapters and primers using cutadapt with the options –pair-adapters which will pair every R1 with its corresponding R2 adapter and –discard-untrimmed which will remove read pairs with missing adapters and –action=trim to remove the adapter.
(2) Create_meta: This takes the input folder of trimmed fastq files and creates an input sample list for the next process by finding the correct sample pattern (--pattern_fw *R1.trimmed.fastq.gz and --pattern_rv *R2.trimmed.fastq.gz)
(3) Run_dada2R: This process performs the dada2 based analysis of the trimmed files. The following parameters were used:

(a) class: "parasite" as we looked at Plasmodium samples
(b) maxEE: "5,5" this represents the maximum number of expected errors allowed
(c) trim_right: "10,10”: we noticed that the amplicon sequencing data occasionally lost quality in the last 4-10 reads requiring this option to cut off 10bp from the right
(d) min_length: 30: discard reads that are shorter than 30bp
(e) truncQ: "5,5” reads will be truncated as soon at the first base phred quality score is below 5
(f) max_consist: 10 The maximum number of steps when selfConsist=TRUE
(g) omegaA: 1e-120: treshold for significantly overabundance
(h) justconcat: 0: turn off concatenation instead of merging
(i) platform: "PE”: set the platform to paired-end reads
(j) trimqv: 15

(4) post_process: custom script that turns the dada2 output seqtab table into an ASV (Amplicon Sequence Variant) Table as well as and ASVSeqs fasta file using the (3) fasta file
(5) asv_to_cigar: this custom script transforms the ASVs into CIGAR (Concise Idiosyncratic Gapped Alignment Report) strings.
